# Accounting for the long-distance transmission route: an epidemiological model of airborne disease transmission in hospitals

**DOI:** 10.64898/2025.12.12.25341907

**Authors:** Olivier Gaufrès, Quentin J. Leclerc, Julien Derdevet, George Shirreff, Solen Kernéis, Lulla Opatowski, Laura Temime, Maylis Layan

## Abstract

Nosocomial transmission of respiratory infections poses a major threat to patient safety, while also affecting healthcare workers’ (HCW) health, generating substantial costs for hospitals. These infections spread through both close-proximity interactions at short distances, and via aerosols that remain suspended in the air, enabling long-range transmission. The relative contribution of each transmission route is pathogen-dependent, and evidence to distinguish them remains scarce. Here, we propose an agent-based stochastic model of respiratory pathogen transmission in a hospital ward that integrates both transmission routes together with contact patterns and individual movements. After informing our model with real close-proximity interaction data collected in two French intensive care units, we simulate a range of combinations of short- and long-range transmission levels. We select parameter values that keep overall ward transmission intensity stable across combinations. We find that the predominance of one route over another has little effect on overall outbreak dynamics, though the impact on individuals varies markedly. Patients are mostly at risk of short-range transmission from HCWs, while HCWs are mostly affected by whichever route is predominant. This directly influences intervention effectiveness. Universal masking emerges as the most effective strategy, reducing both transmission routes. Its stringency can be relaxed with limited loss of effectiveness when combined with ventilation in relevant rooms. Importantly, intervention ranking remains robust across parameter values, as confirmed by a sensitivity analysis. This new model highlights the importance of explicitly considering physical mechanisms of transmission, and the need for interventions that remain effective irrespective of pathogen characteristics and ward organization.

## Introduction

Hospitals are high-risk environments for the transmission of respiratory pathogens. Rapid turnover of patient admissions, patient frailty, enclosed spaces, recurrent lack of adequate ventilation systems (1,2), and frequent close-proximity contacts between patients and healthcare workers (HCWs) constitute the basic ingredients for respiratory outbreaks (3). Such local outbreaks pose a significant threat to patient safety, but also increase the occupational risk for HCWs. Overall, healthcare-acquired respiratory infections, including flu, COVID-19, or pneumonia caused by *Mycobacterium tuberculosis* lead to high costs for hospitals (3,4). To combat them, continuous surveillance and reactive infection prevention and control (IPC) interventions are key, particularly during seasonal epidemics.

Many microorganisms (bacteria, viruses, fungi) transmit through the air, meaning that they are transmitted by respiratory particles of varying size exhaled by infectious individuals. The understanding of the mechanisms underlying airborne transmission has advanced significantly during the SARS-CoV-2 pandemic. This process can be divided into two routes: the short-range route and the long-distance route (5–7). The short-range route corresponds to transmission during close-proximity interactions between an infectious individual and a susceptible one. This includes transmission by large and fine aerosols, direct physical contacts and indirect contacts through fomites. Conversely, long-distance transmission occurs either through exposure to pathogen-containing aerosols that remain suspended in the air, or through fomites previously deposited on environmental surfaces. The long-range route is characteristic of indoor spaces. The relative contribution of the short- and long-range routes presumably varies depending on the pathogen, which would have direct implications in terms of prevention and control efforts. For example, *Mycobacterium tuberculosis* is often considered as a pathogen mostly transmitting at long distances due to its high stability in the air (8), while influenza viruses are thought to transmit during close contacts, outside of specific circumstances involving aerosol-generating procedures (9). Further research is required to better characterise pathogen-specific transmission routes.

Mathematical models are powerful tools to describe the transmission of nosocomial respiratory infections, understand its key drivers, and evaluate control strategies (10–12). The fields of biophysics and epidemiology have both contributed to the modelling of airborne pathogen transmission in healthcare settings. Biophysical models describe the spread and stability in the air of exhaled pathogen-containing aerosols to assess the risk of infection, generally in relation to air ventilation (11). Some are derived from the simple Wells-Riley model (13), while others make use of highly detailed and parameter-rich computational fluid dynamics (CFD) approaches (11), that generally require strong assumptions on intrinsic pathogen characteristics and assess infection risk over short time periods (typically a few hours). At the other end of the spectrum, epidemiological models have historically focused on transmission during close-proximity interactions, and don’t explicitly allow for transmission over longer ranges. Within healthcare settings, epidemiologists generally prefer stochastic models due to the small size of the involved population, as well as agent-based models that can easily integrate inter-individual heterogeneities and complex temporal networks of contacts (14,15), determinant factors for the design of effective interventions (15–17). Between these two ends, some studies have attempted to combine both transmission routes, linking the fine-grain temporal scale of rapid biophysical processes with the coarser temporal scale of inter-individual transmission. However, most assume homogeneous mixing between patients and healthcare workers (18,19), although contacts are known to be highly structured (14,20). Interestingly, two studies have tried to evaluate the relative contribution of the short-range, long-range and fomite routes in hospital wards using data from nosocomial SARS and MERS-CoV outbreaks (18,19). However, in these studies, many parameters were calibrated with values from the literature without exploring their impact in a sensitivity analysis, and the grid search approach used for parameter estimation does not guarantee an optimal exploration of the parameter space, limiting the interpretability of their results. A recent study integrated a CFD component into a transmission chain reconstruction model to evaluate the role of long-range transmission in SARS-CoV-2 nosocomial circulation (21). They calibrated their model over more than two years of data but had no access to HCW information, and thereby overlooked a major actor of nosocomial transmission.

From an epidemiological and public health perspective, a thorough understanding of the respective roles of the short- and long-range routes in nosocomial respiratory outbreaks is essential to develop effective control strategies (6). Here, we propose a novel dynamic epidemiological model that describes the transmission of a range of respiratory pathogens in a hospital ward. After presenting the model that accounts for complex temporal inter-individual contact networks, individual movements between ward rooms, and exposure to pathogen-laden aerosols suspended in the indoor air, we apply it to records of close-proximity interactions in two French intensive care units (ICUs). Then, we explore with our simulation platform how varying the levels of the short- and long-range transmission routes impacts epidemic dynamics, using SARS-CoV-2 as a case study. Finally, we assess the effectiveness of IPC interventions, including hand hygiene, mask wearing, and air ventilation, to identify the best strategy for each contribution level of the two transmission routes.

## Results

### Overview of the transmission model and the simulation platform

We describe the circulation of a respiratory pathogen among patients and HCWs using a stochastic discrete-time model in which individuals can be susceptible, exposed, infectious, or recovered (Fig 1). Susceptible individuals get infected either during contacts with infectious individuals or following the inhalation of infectious aerosols in the rooms they visit. These two transmission routes feed into the global force of infection. Infectious aerosols within a room are assumed to be well-mixed. The dynamics of their concentration is governed by an exponential decay due to pathogen inactivation and a linear increase when infectious individuals are present in the room. This model relies on contacts between individuals and on movements between rooms of the hospital ward. Since contact data, when available, typically cover only a few days in healthcare settings and exact individual movements are not known (20,22–26), we propose a procedure that generates synthetic temporal networks from observational studies using a published algorithm (27) and reconstructs movements based on the category (i.e. patient, HCW) and the contacts of the individual using deterministic decision trees (SI Appendix, Fig S3). Here, we apply our simulation procedure to SARS-CoV-2 for illustrative purposes. Our choice is motivated by the availability of estimates for model parameters in the literature.

**Fig 1.**
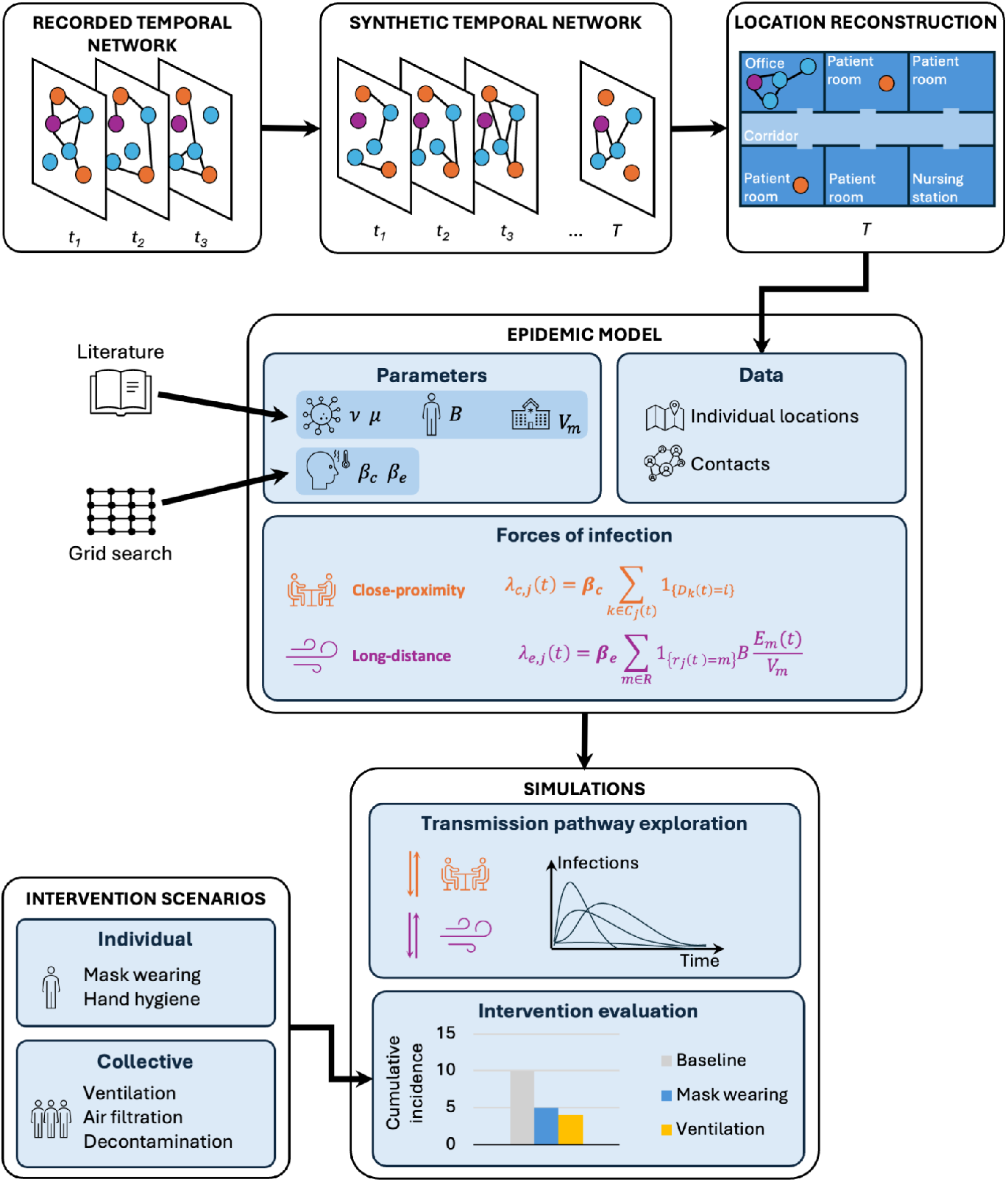
Modelling framework. We developed a nosocomial transmission model of a respiratory pathogen that accounts for the close-proximity and long-distance transmission routes. Individual locations and interindividual contacts are informed by synthetic temporal networks reconstructed from contacts recorded in two French ICUs. Other parameter values (aerosol excretion rate *v*, aerosol inactivation rate *μ*, breathing rate *B*, and room volumes *V*_*m*_) are set based on estimates from the literature. We tested five (*β*_*c*_, *β*_*e*_) value pairs corresponding to transmission pathways with varying contribution levels of the two transmission routes. Using simulations, we compared the impact of the transmission pathways on outbreak dynamics over a 90-day period, and we evaluated the performance of individual and collective infection prevention strategies.

### Application to close-proximity interactions in two intensive care units

We apply our model and simulation platform to close-proximity interactions recorded in two adult ICUs during the Nods-Cov-2 study (14). Briefly, the Nods-Cov-2 study took place between April and June 2020 during the first and second waves of the SARS-CoV-2 pandemic. It aimed to collect close-proximity interactions using wearable proximity sensors over a 36-hour period across different wards and hospitals in France. Given the limited duration of the records, we augment the number of patients to reproduce realistic hospitalization stay durations over a 90-day period. We then generate a synthetic temporal network over the 90-day period for each ICU. These synthetic networks reproduce well the number of contacts per hour and average contact durations (S1 Fig), allowing to simulate individual movements that are globally consistent with the observed data (Fig 2A and Appendix S2). Using this reconstructed location data, our model is able to generate aerosol dynamics in the different rooms through the movements of infectious individuals (Fig 2B). An example of movement reconstruction at the ward level is available in S1 Movie. We are also able to implement interventions that reduce aerosol concentrations despite the presence of infectious individuals (Fig 2C).

**Fig 2.**
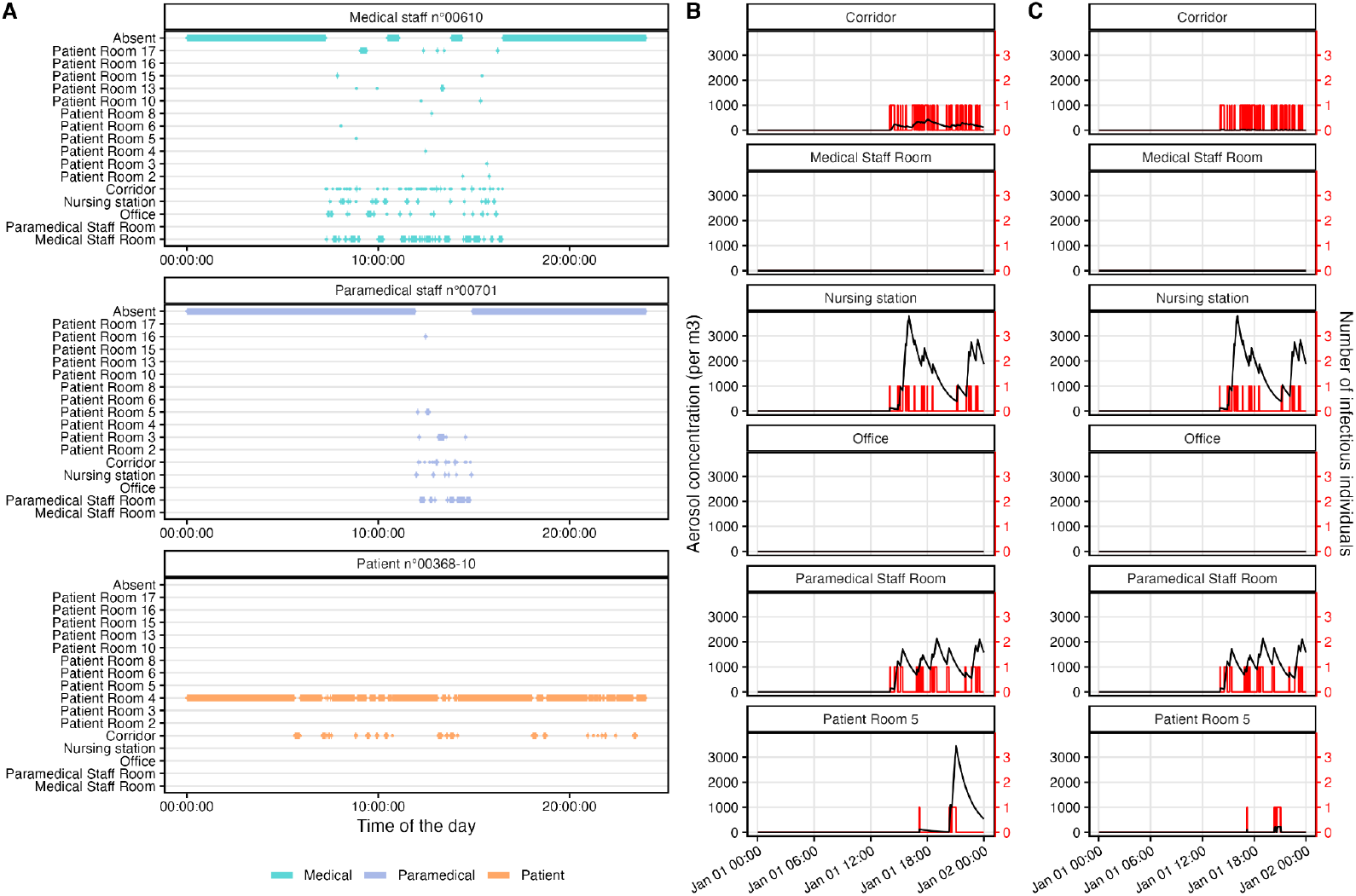
Illustration of simulation outputs. (**A**) Example of movements over one day for three randomly selected individuals among the medical staff, paramedical staff, and patients in ICU1. (**B-C**) Example of simulated aerosol concentrations (in black) in a subset of rooms in ICU1 without intervention (**B**) and with improved ventilation in rooms accessible to patients (**C**). The number of infectious individuals present in the room is depicted in red at each time step. Aerosol concentration increases when infectious individuals are present and decreases otherwise due to pathogen inactivation. When increasing the ventilation to 8 air changes per hour (ACH) in rooms accessible to patients (corridor and patient room n°5), the concentration of aerosols is much lower than in the baseline scenario with no intervention.

### Transmission pathways affect patients and healthcare workers differently

The relative contributions of the long- and short-range transmission routes to nosocomial respiratory epidemics remain poorly characterized and very likely depend on the pathogen. Here, we explore how the relative importance of one or the other transmission route modulates transmission dynamics and changes affected populations. We define five pathways, detailed in Table 1, that give more importance to one or the other transmission route, while maintaining similar global transmission intensity within the ICU (Fig 3B; SI Appendix, Fig S5). Overall, the pathways that we have defined allow us to generate epidemics of similar size within a given ward, with variations due to the stochasticity of the model (Fig 3B), while giving different levels of contribution for the two routes as depicted in Fig 3C. Importantly, we predict a median secondary attack rate (SAR) over 90 days of 49% (IQR: 7%) in ICU1 and 22% (IQR: 25%) in ICU2 (Fig 3B) which is credible for SARS-CoV-2 (28–31). The large difference between the two ICUs is due to the differences in their network properties (Fig 3A; S2 Fig) and time spent by individual categories in the different wards of the ICU (SI Appendix, S4 Fig). The lower overall transmission intensity in ICU2 leads to a lower difference between the relative contributions of the two transmission routes compared to ICU1 (Fig 3C). Other metrics that characterize the general transmission dynamics are also conserved across transmission pathways within an ICU. This is the case of the extinction probability (Fig 4A), the epidemic duration (Fig 4B), and the time to the peak (Fig 4C). However, stratifying SARs by individual categories reveals that patients are less at risk (26%, IQR: 8% in ICU1 and 8%, IQR: 11% in ICU2) than HCWs (76%, IQR: 10% in ICU1 and 50%, IQR: 58% in ICU2). This is due to the short hospitalization stay of patients and to HCWs having more contacts than patients (S1A Fig). Overall, HCWs are responsible for most of the transmission events at short distances (S6 Fig) and they are more at risk of acquisition due to persistent aerosols when long-distance transmission is predominant (Fig 4D). Patients, on the other hand, generally get infected through the short-distance route (Fig 4D) mostly during their interactions with HCWs (S6 Fig), irrespective of the relative importance of this transmission route compared to the long-distance one. This can be explained by the fact that patients spend most of their time in their room, thereby being mainly exposed to the aerosols in their room, and they mostly have contacts with HCWs rather than other patients.

**Table 1.** Parameter values used to parameterize the epidemic simulation model. The simulation model is parametrized using values taken from the literature, except for*β*_*c*_ and *β*_*e*_ that we calibrate using a grid search approach. We have retained five value pairs for (*β*_*c*_, *β*_*e*_) to explore transmission pathways of varying contribution levels of the long-distance and close-proximity transmission routes. We have chosen these specific value pairs because they lead to similar secondary attack rates on a given contact network. Importantly, we inform the inactivation rate of aerosols using the early median estimates of SARS-CoV-1 and SARS-CoV-2 half-lives from Van Doremalen et al. (2020) that lie between 1.1 and 1.2 hours. If we consider that the air of a room is completely renewed when the quantity of aerosols has decreased by more than 99%, then a half-life of 1.1 hours is equivalent to an air change per hour (ACH) of 0.13.

| Parameter | Description | Value |  |  |  |  | Unit | Reference |
| --- | --- | --- | --- | --- | --- | --- | --- | --- |
|  |  | Pathway 1 | Pathway 2 | Pathway 3 | Pathway 4 | Pathway 5 |  |  |
| $\beta_c$ | Close-proximity transmission rate | 0.75 | 1.00 | 1.25 | 1.50 | 1.75 | No. of new cases per contact per day | Grid search |
| $\beta_e$ | Long-distance acquisition rate | 1/45 | 1/60 | 1/70 | 1/100 | 1/150 | No. of new cases per aerosol per day | Grid search |
| $\nu$ | Exhalation rate of infectious aerosols | 1.8e5 | | | | | RNA copies per 30 min | Lai et al., 2024 |
| $B$ | Breathing rate | 0.006 | | | | | m<sup>3</sup> air/min | Pleil et al., 2022 |
| $V^k$ | Volume of room $k$ | Single room: 50<br>Double room: 75<br>Medical staff room: 75<br>Paramedical staff room: 156.25 (ICU1) and 100 (ICU2)<br>Nursing station: 96.02 (ICU1) and 50 (ICU2)<br>Medical office: 75<br>Corridor: 757.50 (ICU1) and 550 (ICU2) | | | | | m<sup>3</sup> | ICU1 and ICU2 configurations |
| $\mu$ | Pathogen-laden aerosol removal due to pathogen inactivation | 0.63 (1.1) | | | | | Rate per hour (half life in hour) | van Doremalen et al., 2020 |
| $\Delta T_{incubation}$ | Incubation period | $\Gamma(mean = 4.07, sd = 2.12)$ | | | | | day | Galmiche et al., 2023 |
| $\Delta T_{infectious}$ | Infectious period | U(3,7) | | | | | day | Hakki et al., 2022 |

**Fig 3.**
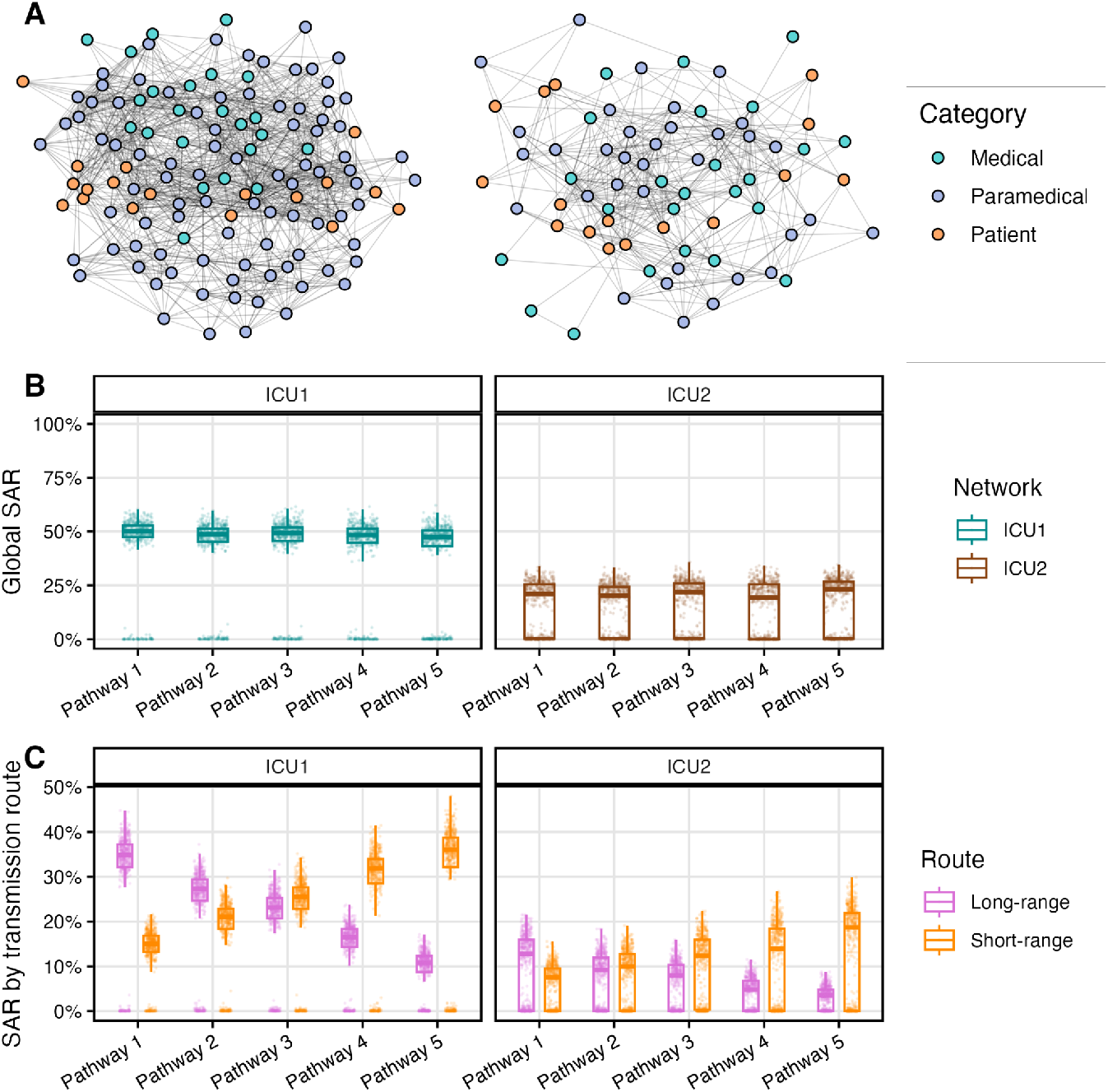
Implementation of the five transmission pathways on two close-proximity contact networks. (**A**) Close-proximity contact network of two French adult ICUs, ICU1 a large ICU and ICU2 a smaller ICU. Colored vertices correspond to individuals with medical staff in blue, paramedical staff in purple, and patients in orange. Two vertices are linked by an edge if they have at least one contact record in the Nods-Cov-2 study. (**B**) Secondary attack rates (SAR) across transmission pathways for *n*=500 simulations, over 90 days. (**C**) SAR stratified by transmission route, over 90 days. Pathway 1: *β*_*e*_ = 1/45 new cases per infectious aerosol per day and*β*_*c*_ = 0.75 new case per contact per day; Pathway 2: *β*_*e*_ = 1/60 and *β*_*c*_ = 1.00; Pathway 3: *β*_*e*_ = 1/70 and *β*_*c*_ = 1.25; Pathway 4: *β*_*e*_ = 1/100 and *β*_*c*_ = 1.50; and Pathway 5: *β*_*e*_ = 1/150 and *β*_*c*_ = 1.75.

**Fig 4.**
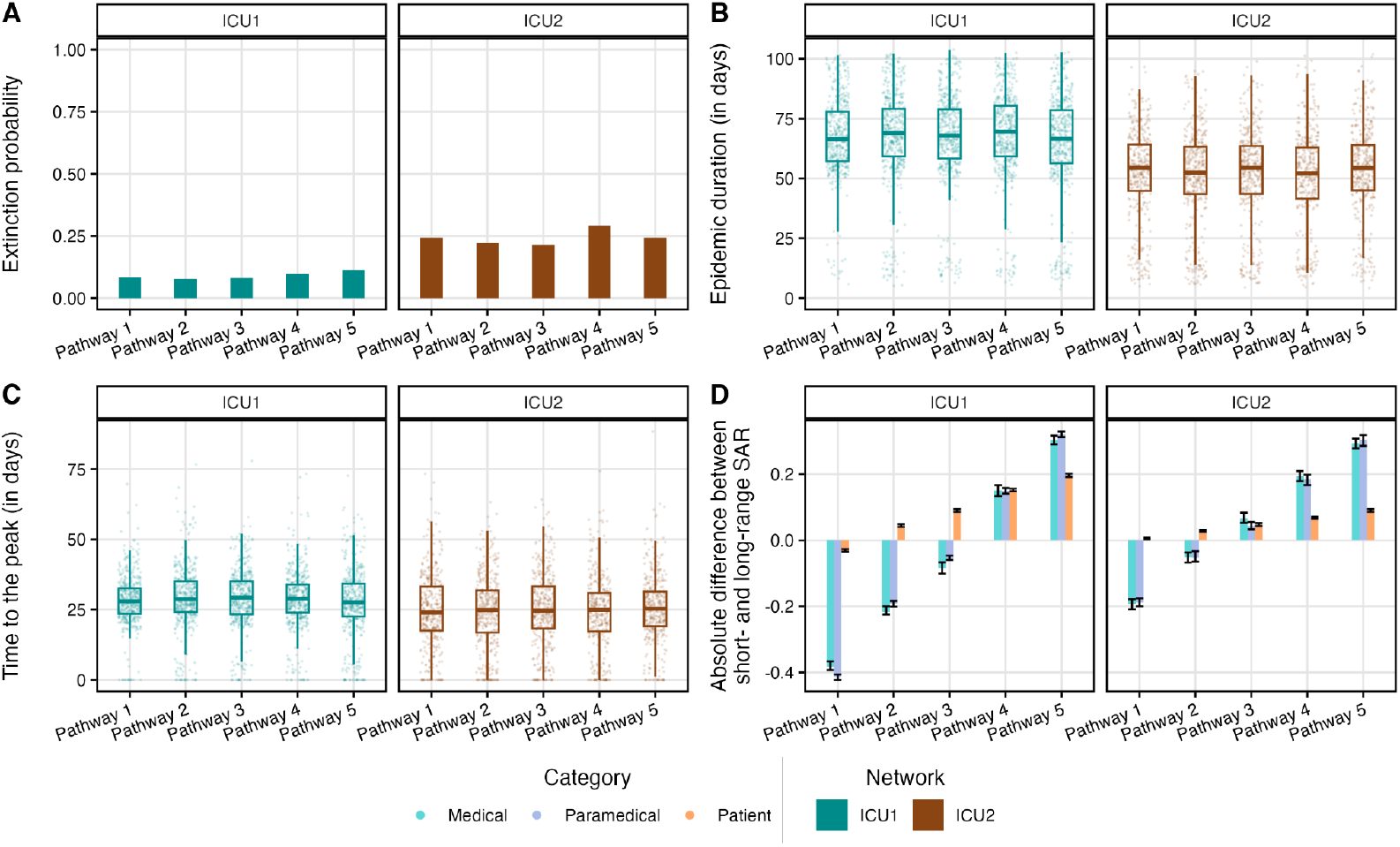
Characterisation of nosocomial transmission dynamics across transmission pathways. We report three metrics of overall transmission dynamics for *n*=500 simulations per transmission pathways and close-proximity contact network: the extinction probability (**A**), the epidemic duration in days (**B**), and the time to the peak in days (**C**). We also report the estimated absolute difference (pseudo-median) from Wilcoxon tests between the SAR related to the close-proximity route and the SAR related to the long-distance route per individual category (**D**). Pathway 1: *β*_*e*_ = 1/45 new cases per infectious aerosol per day and *β*_*c*_ = 0.75 new case per contact per day; Pathway 2: *β*_*e*_ = 1/60 and *β*_*c*_ = 1.00; Pathway 3: *β*_*e*_ = 1/70 and *β*_*c*_ = 1.25; Pathway 4: *β*_*e*_ = 1/100 and *β*_*c*_ = 1.50; and Pathway 5: *β*_*e*_ = 1/150 and *β*_*c*_ = 1.75.

### Control strategies perform differently across transmission pathways

The pattern of pathogen circulation is shaped by the transmission pathway which may, in turn, affect the effectiveness of control strategies. We investigate this interplay by testing seven intervention scenarios detailed in Table 2 that are either individual (universal or targeted mask wearing, improved hand hygiene among HCWs), collective (ventilation), or mix both types of interventions. Mask wearing reduces short-range transmission and exhalation of infectious aerosols, hand hygiene reduces transmission from HCWs to patients during close-proximity interactions, and ventilation increases the inactivation rate of infectious aerosols. Intervention efficacy on model parameters is set using estimates from the literature.

**Table 2.** Description of intervention scenarios. We tested seven intervention scenarios that we compared to the baseline scenario with no intervention. While hand hygiene targeted the close-proximity route and ventilation targeted the long-distance route, wearing masks and mixed interventions limited both long-distance and close-proximity routes.

| Intervention | Type | Parameter values | References |
| --- | --- | --- | --- |
| None | - | - | - |
| Improved hand hygiene among HCWs | Individual | 17% ↘ $\beta_{c,HCW \rightarrow patient}$ | Aiello et al., 2008<br>Jefferson et al., 2008<br>Ross et al., 2023 |
| Targeted mask wearing | Individual | 70% ↘ $\beta_{c,HCW \leftrightarrow patient}$<br>70% ↘ $\beta_{c,patient \leftrightarrow patient}$<br>94% ↘ $\nu$ | Offeddu et al., 2017<br>SF2H report |
| Universal mask wearing | Individual | 70% ↘ $\beta_c$<br>94% ↘ $\nu$ | Offeddu et al., 2017<br>SF2H report |
| Patient room ventilation (Patient rooms + corridor) | Collective | ↗ $\mu$ to an ACH of 8 | Zhang et al., 2023 |
| HCW room ventilation (Medical office + Nursing station + Paramedical staff room + Medical staff room) | Collective | ↗ $\mu$ to an ACH of 8 | Zhang et al., 2023 |
| Targeted mask wearing + Patient room ventilation | Collective + Individual | 70% ↘ $\beta_{c,HCW \leftrightarrow patient}$<br>70% ↘ $\beta_{c,patient \leftrightarrow patient}$<br>↗ $\mu$ to an ACH of 8<br>94% ↘ $\nu$ | - |
| Targeted mask wearing + HCW room ventilation | Collective + Individual | 70% ↘ $\beta_{c,HCW \leftrightarrow patient}$<br>70% ↘ $\beta_{c,patient \leftrightarrow patient}$<br>↗ $\mu$ to an ACH of 8<br>94% ↘ $\nu$ | - |

Intervention effectiveness is quantified by comparing the 90-day cumulative incidence in patients to the baseline scenario without intervention. In Fig 5A, we present intervention effectiveness in patients by ICU and transmission pathway. Despite higher variability in the predicted impact of interventions in ICU2— likely because of its lower overall transmission level giving more importance to stochastic variations—the overall performance of interventions is remarkably similar in both ICUs. The effectiveness of hand hygiene and ventilation depends on the predominant transmission route. These interventions are more effective when they target the predominant route of the transmission pathway (Pathway 5 for hand hygiene and Pathway 1 for ventilation, Fig 5A). In contrast, mask wearing, be it universal or designed to primarily protect patients (“targeted masking”), works equivalently across pathways.

**Fig 5.**
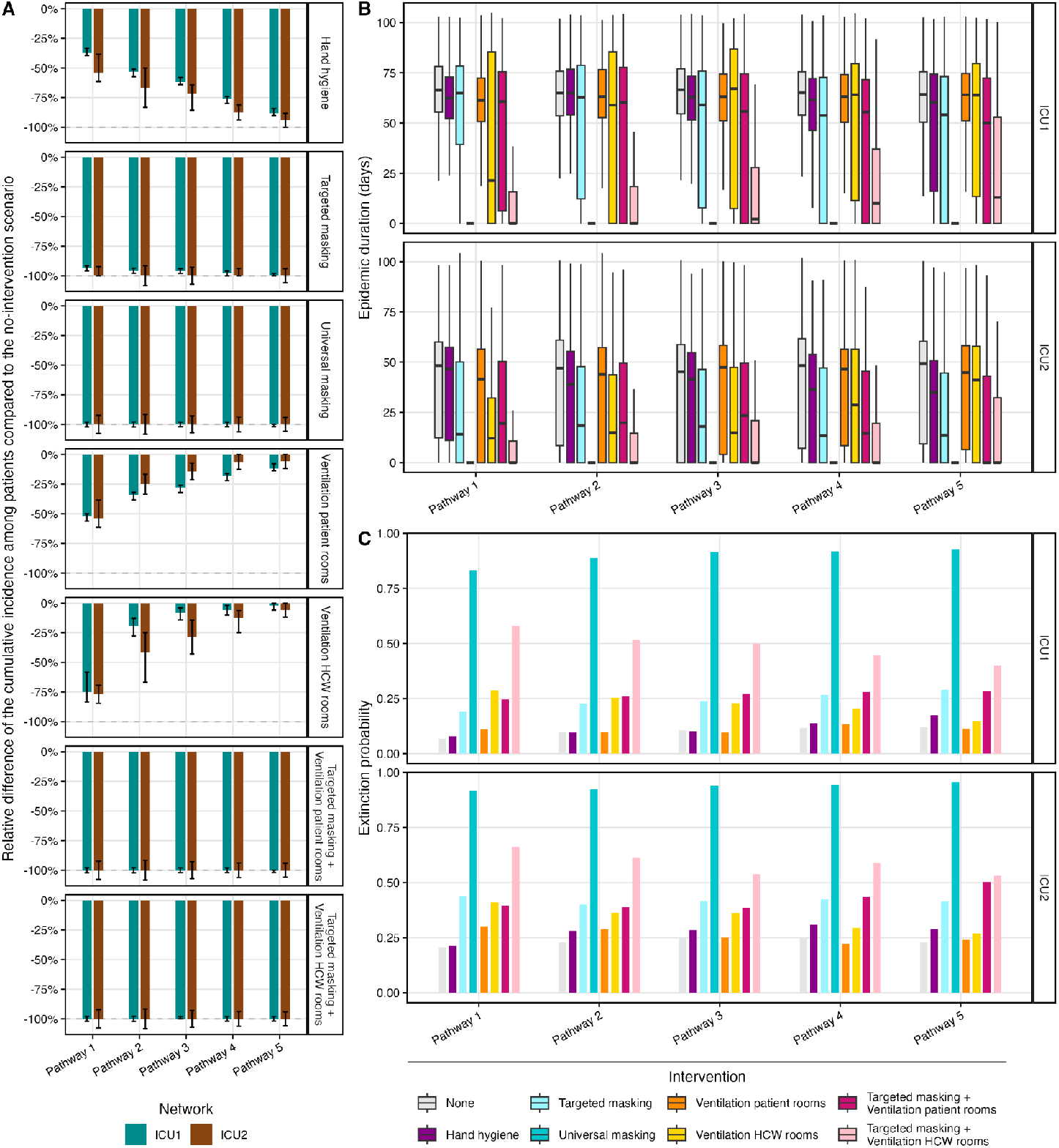
Impacts of control interventions on secondary attack rates, epidemic duration, and extinction probability across transmission pathways. (**A**) Relative difference of the 90-day cumulative incidence among patients between the intervention scenario and the baseline scenario (with no intervention). Negative values indicate a reduction in the cumulative incidence when an intervention is in place. Relative difference estimates and error bars are derived from the pseudo-median and the 95% CI of Wilcoxon tests, respectively. Reductions stronger than 100% are explained by the definition of the metric: the median of the cumulative incidence in the baseline scenario used as the reference can be higher than the 95% CI bounds of the difference. (**B**) Distribution of epidemic durations in the baseline and intervention scenarios. (**C**) Extinction probability in the baseline and intervention scenarios. Pathway 1: *β*_*e*_ = 1/45 new cases per infectious aerosol per day and *β*_*c*_ = 0.75 new case per contact per day; Pathway 2: *β*_*e*_ = 1/60 and *β*_*c*_ = 1.00; Pathway 3: *β*_*e*_ = 1/70 and *β*_*c*_ = 1.25; Pathway 4: *β*_*e*_ = 1/100 and *β*_*c*_ = 1.50; and Pathway 5: *β*_*e*_ = 1/150 and *β*_*c*_ = 1.75.

**Fig 6.**
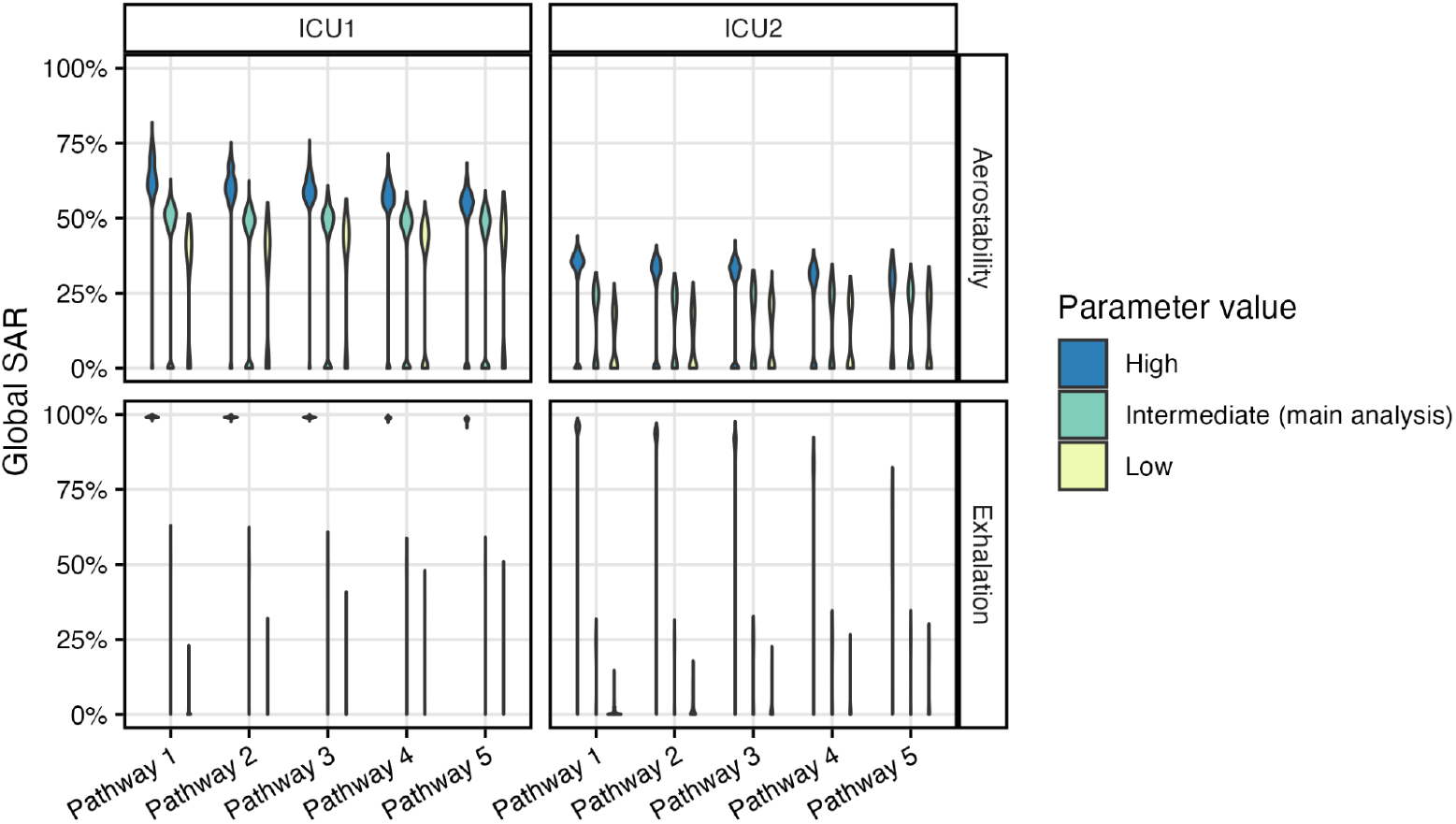
Role of the magnitude of the aerostability and the exhalation rate of infectious aerosols on overall epidemic dynamics. We tested low, intermediate, and high values of aerostability and exhalation rate of infectious aerosols (in rows) in a univariate sensitivity analysis across the two ICU networks and the five transmission pathways. Parameter values are detailed in Table 3. Violins represent the distribution of the global SAR for each condition. Pathway 1: *β*_*e*_= 1/45 new cases per infectious aerosol per day and *β*_*c*_ = 0.75 new case per contact per day; Pathway 2: *β*_*e*_ = 1/60 and *β*_*c*_ = 1.00; Pathway 3: *β*_*e*_ = 1/70 and *β*_*c*_ = 1.25; Pathway 4: *β*_*e*_ = 1/100 and *β*_*c*_ = 1.50; and Pathway 5: *β*_*e*_ = 1/150 and *β*_*c*_ = 1.75.

Universal mask wearing surpasses all the other interventions; it reduces in all transmission pathways up to 100% of the incidence in patients (Fig 5A) and HCWs (SI Appendix, Fig S7) by preventing transmission events at the very start of the outbreak. This is indeed the intervention with the highest extinction probability in all transmission pathways (Fig 5C). Although targeted masking effectively prevents infection in patients (Fig 5A), transmission still occurs among HCWs (SI Appendix, S7 Fig). In the case of ventilation, targeting rooms that are not accessible to patients is more effective in terms of patient incidence than targeting patient rooms (Fig 5A). Indeed, targeting rooms that are not accessible to patients reduces infections in HCWs, that is to say the first source of infection for patients. When combined with targeted masking, ventilation is as effective as universal masking in terms of patient incidence (Fig 5A), and in terms of HCW incidence as well (SI Appendix, S7 Fig), but only when ventilation targets HCW rooms. Overall, coupling targeted masking with ventilation in HCW rooms reduces acquisition among HCWs as well as transmission from HCWs to patients (SI Appendix, S9 Fig). This mixed intervention is thereby almost as effective as universal masking, although its efficacy tends to decrease for pathways where the short distance route is predominant (Pathway 5; SI Appendix, S8 Fig).

### Univariate sensitivity analysis

Biological characteristics of respiratory pathogens and intervention effectiveness against infection are difficult to quantify precisely, both in controlled experiments and in observational studies. The parameter values that we used in the main analysis (Table 1) do not reflect the full range of possible values for each parameter. To assess the impact of this uncertainty, we evaluate changes in nosocomial dynamics when using upper and lower bounds of pathogen aerostability, pathogen exhalation, and intervention effectiveness (Table 3). As expected, higher aerostability or higher exhalation rate of infectious aerosols increase the global SAR. Concerning interventions, increasing or decreasing their effectiveness does not modulate overall dynamics (SI Appendix, S10) and little affects the ranking of the most effective interventions (SI Appendix, S11-13 Figs).

**Table 3.**
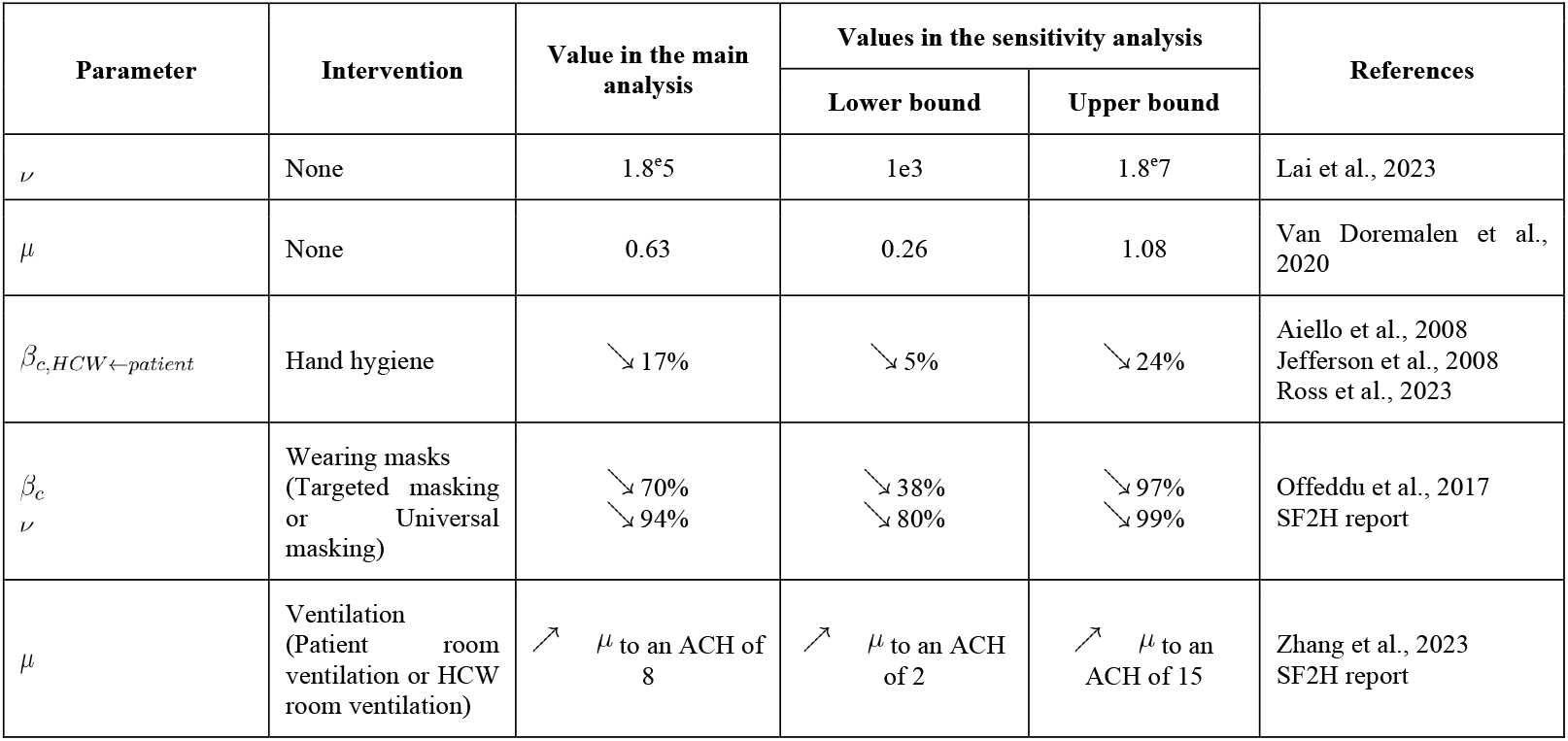
Description of the univariate sensitivity analyses.

## Discussion

In our study, we propose and test a novel model and simulation framework of nosocomial outbreaks that integrate short- and long-distance transmission routes, while reconstructing proximities and individual locations from time-resolved close-proximity interactions data. This framework is applicable to any ICU. In our case study, we show that varying the contribution levels of the two transmission routes changes the epidemiology in patients and HCWs, highlighting the need for adapted interventions.

Using a grid search approach, we have selected five transmission pathways giving more weight to one transmission route or the other while maintaining similar circulation intensity in the ward. Although we used SARS-CoV-2 for illustration purposes, the model can easily be extended to other airborne pathogens. Besides, our general conclusions are likely to be applicable to other respiratory viruses, such as the respiratory syncytial virus or influenza viruses, that have similar transmission characteristics as SARS-CoV-2 (9,32). Regardless of the predominant transmission route, our results suggest that patients in ICUs are mostly infected during close-proximity interactions with HCWs, highlighting the need to control transmission among HCWs. These results are very likely to vary in other healthcare settings, such as nursing homes, long-term care facilities or other acute-care wards, where patients have more interactions with other patients, as well as visitors, and have access to other rooms (20,23). For instance, in two recent studies on SARS-CoV-2 circulation in hospital wards, the authors found that HCWs play a negligible role in patient infection (21,33). This could be due to the organization of the wards with multi-bed rooms. Conversely, in another modelling study exploiting national-level data, the authors showed that intermittent SARS-CoV-2 carriage by HCWs constitute the primary source of infection for patients (34) which is consistent with our simulations. These contrasting results demonstrate the importance of data on HCWs in identifying the sources of infection for patients, and yet they are often the missing piece of the puzzle. This also has implications in the design of control strategies as we show that improving ventilation in rooms that are not accessible to patients cuts down transmission among HCWs, thus indirectly reducing the incidence in patients. This confirms that interventions should not only target patients, but also HCWs, as suggested by Evans et al. (34).

In the present study, we highlight how differences in transmission mechanisms call for mixed control interventions. If we design intervention strategies that solely target one transmission route, which is the case for hand hygiene and ventilation, we face a strong decreasing effectiveness for pathogens that transmit more by the other route. The best interventions are the ones that curb both transmission routes. In our simulations, masks appear as the most effective strategy, as they may help control transmission in all transmission pathways and contact networks. However, we assume here universal masking from the symptom onset of the index case. Such drastic rules would face compliance reduction over time. Thus, the best intervention should reduce both transmission routes, be effective for a wide range of contact networks, be considered acceptable by patients and HCWs, and should not increase the workload of HCWs. Collective interventions such as ventilation are convenient and alleviate workload for HCWs. They should be taken into account at the design stage of construction projects. However, they require funding (2) and have a limited impact on short-distance transmission as exemplified here. They should therefore be combined with physical interventions that are reinforced during epidemic seasons. This is exemplified here by the intervention that couples ventilation in HCW rooms and targeted masking. This strategy is effective to reduce patient incidence and HCW incidence in most transmission pathways, although the effectiveness in HCWs decreases when the short-range route predominates. Importantly, we have tested different levels of mask effectiveness in the sensitivity analysis covering surgical masks and respirators. We do not observe a strong difference in the overall impact on epidemic dynamics between surgical masks (lower bound in the sensitivity analysis) and respirators (upper bound in the sensitivity analysis) which is consistent with the literature on respiratory viruses (35,36). Finally, short-distance transmission in our model is a bundle of (in)direct physical transmission and transmission through aerosol deposition and inhalation. The relative importances of these different routes at short distances may also vary across pathogens which highlights the importance of other physical interventions, such as wearing gowns, that have proved effective in reducing respiratory viruses transmission (37).

We acknowledge several limitations in our modelling approach. First, we opted for a straightforward room-by-room modelling approach of aerosol dynamics. Consequently, our results on intervention effectiveness should be interpreted in light of two key model assumptions: *i*) the immediate homogeneous mixing of aerosols within each room and *ii*) the absence of aerosol diffusion across rooms. While the first assumption is justifiable given that exact movements within rooms are unknown and would be difficult to collect related to acceptability issues, the second assumption probably leads to the underestimation of the contribution of the long-distance route in overall transmission dynamics. Refining these dynamics necessitates the use of CFD models. This has been initiated in a recent study where the authors reconstructed exact transmission events between patients in a data-driven modelling approach that combines probabilistic transmission chain reconstruction and exposure to infectious aerosols using CFD modelling (21). However, this study did not account for HCWs in transmission dynamics nor for individual movement trajectories between rooms. By complexifying aerosol dynamics, we could account for important physical and biological mechanisms, including among others the role of thermal plumes on aerosol transport (38), or the role of relative humidity (39) or carbon dioxide concentration (40) on the aerostability of pathogens. This would also allow us to explicitly model the mechanisms of different air cleaning technologies (ventilation, filtration, germicidal UVs etc…) whose effectiveness remains poorly quantified (41).

Another major limitation is that we consider biological, biophysical, and epidemiological processes as constant over time and homogeneous across individuals. A wide body of literature notably highlighted the variability of infectiousness over the course of the infectious period and across individuals (42), the variability in infectious aerosol generation depending on individual heterogeneity (43–46), symptoms or aerosol-generating procedures (47) (although this is still debated (48,49)), and variations in breathing rates related to medical conditions (50) and medical procedures (51). Nevertheless, our aim was to develop a generic model applicable to multiple pathogens. Importantly, we do not allow for longer hospitalization stays when patients get infected, a common consequence that generates high costs for hospitals and may increase patient load (52). Such consideration is important notably to evaluate costs. Another limitation relates to the absence of transient hand carriage in HCWs in our model, although this has been shown to be an important route for viruses (34), and even more so for bacteria (53). Accounting for this route would increase the impact of hand hygiene and material disinfection on transmission dynamics in our simulations. In addition, we did not consider baseline immunity levels in the population related to previous vaccination or infection. Vaccination is often promoted among HCWs, so introducing a baseline vaccination coverage in our simulations could decrease the role of HCWs in transmission dynamics. However, vaccine coverage and availability vary across countries, pathogens and healthcare roles (54). For instance, flu vaccination coverage lies around 20% (55), while it reaches approximately 77% for COVID-19 (56). The choice of prior immunity prevalence and prior immunity protection against infection depends on the pathogen, the subpopulation and the context (pandemic or epidemic) under consideration. Of note, our location reconstruction algorithm is generic and relies on few assumptions, so that it remains flexible enough to apply to different hospitals. However, it is tailored to ICUs and would require adjustments to be generalized to other types of wards (e.g. geriatric wards, long-term care facilities). Finally, the Nods-Cov-2 study took place during the COVID-19 pandemic, when patients were more severe and HCWs more numerous. Therefore, interaction dynamics may not be generalizable outside of pandemic times.

Overall, the nosocomial transmission of respiratory pathogens is a complex phenomenon influenced by multiple biophysical and behavioral factors. We developed a novel model that integrates aerosol dynamics and individual contact data to better understand how short- and long-range transmission routes weigh on patient and HCW safety and identify the key ingredients of an effective intervention. Further efforts should be deployed to quantify the relative contribution of the two routes from observational studies tailored to specific pathogens and settings. Modelling can help design the collection of data in hospitals for these future studies, and quantify the impact of control strategies. Such studies would be instrumental for epidemic and pandemic preparedness in hospitals.

## Materials and Methods

### Nosocomial transmission model of respiratory infections

We formulate the model in discrete-time. For notational brevity, time *t* refers to the time step between *t* and *t* + *dt* with *dt* > 0 and small enough so that only one competing event (e.g. infection and recovery) can happen within a single time step. Four disease statuses are possible: individuals can be susceptible, exposed, infectious, or recovered. The probability *P*^*j*^ (*t*) that a susceptible individual *j* is infected between *t* and *t* + *dt* is given by :

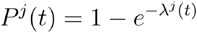

This probability depends on the force of infection *λ*^*j*^ (*t*) that is the sum of the force of infection due to close-proximity interactions with infectious individuals 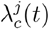, and the force of infection due to pathogen-containing aerosols in suspension in the indoor air 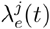.

The force of infection due to close contacts is defined as:

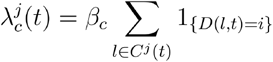

Where *β*_*c*_ is the person-to-person transmission rate during a close-proximity interaction with an infectious individual, *C*^*j*^ (*t*) is the set of individuals in close-proximity interaction with individual *j* at time *t* and *D* (*l, t*) is the disease status of contact *l* (*s*: susceptible, *e*: exposed, *i*: infectious, and *r*: recovered) at time *t*. Therefore, the sum 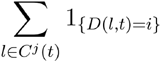 corresponds to the number of infectious individuals in interaction with individual *j* at time *t*.

The force of infection due to long-range airborne transmission 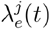 was formulated using a linear dose-response model

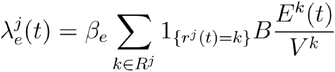

Where *β*_*e*_ is the acquisition probability following exposure to an infectious particle, *R* ^*j*^ is the set of rooms visited by *j* over the study period, 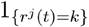 is the indicator function designating the room visited by *j* between *t* and *t* + *dt, B* is the respiratory frequency of the susceptible individual, *E* ^*k*^ (*t*) is the quantity of infectious particles in room *k*, and *V* ^*k*^ is the volume of room *k*. The ratio 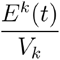 corresponds to the concentration of infectious particles in room *k* at time *t*. When multiplied by the respiratory frequency *B*, we obtain the quantity of inhaled infectious particles by unit of time. The quantity of infectious particles *E* ^*k*^ (*t*) is iteratively defined as:

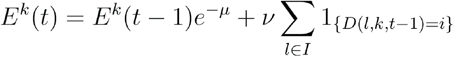

Where the first term corresponds to the first-order exponential decay of infectious particles with inactivation rate *μ*, and the second term represents the exhalation of new infectious aerosols in room *k* with *v* is the exhalation rate of infectious aerosols,*I* is the set of infectious individuals, and *D* (*l, k, t* − 1)the disease status of individual *l*.

Once an individual is infected, we draw its incubation period 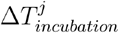 and infectious period 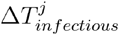 from pre-specified distributions. Thus, if an individual is infected at time *t*, it goes through the following stages:

1. Susceptible from *t*_0_ to *t*
2. Exposed from *t* to 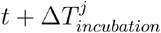
3. Infectious from 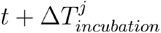 to 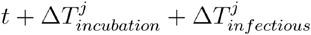
4. Recovered from 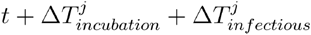 to *t*_*end*_

After each infection event, we sample the source of infection *h* (e.g. the contact with an infection individual or the long-distance route) using the following probability:

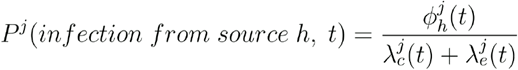

Where 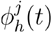 is either 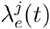 or the force of infection due to the contact with one infectious contact *β*_*c*_. This sampling procedure allows the reconstruction of transmission chains at the end of the epidemic in the case of short-range transmission.

### Location reconstruction algorithm

Our model accounts for exposure to infectious aerosols when individuals visit the different rooms of the ward, data that are rarely available (20,22–26). We propose a location reconstruction algorithm detailed in S2 Appendix that generates the movements for every individual across the rooms of a minimal ward. The configuration of this minimal ward is generic and contains a nursing station, an office for the medical staff, separate rooms for medical and paramedical staff, patient rooms, and a corridor that connects all the rooms. In this algorithm, movements depend on the category of the individual (e.g. patient or HCW) and their contacts.

### Application on SARS-CoV-2 using real close-proximity interaction data from intensive care units

To illustrate the practical utility of our model, we apply it to real world close-proximity interactions data collected during the Nods-Cov-2 study that has been previously described (14,23). Briefly, close-proximity interactions at less than 1.5 meters were recorded every ten seconds between volunteers (including patients, HCWs, visitors, and other staff) with wearable sensors over approximately 36 hours. The study was carried out between April and June 2020 in 17 wards distributed across nine hospitals in France. Here, we focus on the contact data from two adult ICUs located in two different hospitals, denoted ICU1 and ICU2. This choice is motivated by the low number of visitors enrolled (*n*=1 and *n*=0, respectively), the large number of participants (*n*=166 and *n*=92, respectively), and the limited movement of patients within this type of ward, facilitating the reconstruction of their location. Here, we exclude volunteers that are not patients or HCWs (i.e. visitors, administrative staff, logistic staff).

Our aim is to simulate epidemics of respiratory infections over 90 days using realistic ward organizations. Given the limited duration of contact recording in Nods-Cov-2, we extend the contact network using synthetic data that reproduce realistic stay lengths, patient turnover in ICUs, and contacts between individual categories. First, we augment the number of patients by drawing hospital stay durations in a uniform distribution spanning from six to 13 days (57). This allows us to increase the number of patients from 18 to 186 in ICU1, and from 17 to 175 in ICU2 over 90 days. Second, we generate synthetic temporal networks on these new sets of individuals using a published algorithm (27) (Fig 1). In brief, this algorithm uses a linelist of individuals and close-proximity interaction records to stochastically generate the number and duration of interactions for each individual while accounting for the average per-capita contact rate at a given hour of the day and the probability of recurring contacts hour after hour. The algorithm enables contacts to be reconstructed for individuals with no recorded interaction (e.g. augmented patients) by considering that not all close-proximity interactions are observed due to measurement errors. Details on the comparison of the recorded interactions and the synthetic temporal networks are available in S1 Appendix.

### Simulation of nosocomial transmission of SARS-CoV-2

#### Parameter values

We simulate the transmission of a virus after introduction in the ward. Each simulation starts by the introduction of a random index case and runs over 90 days. Parameter values, chosen to illustrate SARS-CoV-2 transmission, are listed in Table 1. Of particular note, room volumes are based on a height of 2.5m and are adapted so that the volume per individual is equivalent at maximum capacity in both ICUs.

All pathogen-related parameters, except *β*_*c*_ and *β*_*e*_, are informed using estimates from the literature on SARS-CoV-2, the airborne pathogen that has been documented in the most detail. For the exhalation rate of infectious aerosols, we use an estimate of the total number of RNA copies exhaled by a COVID-19-positive case during 30 minutes from Lai et al., 2023 (43). This way, we make the assumption that pathogens are homogeneously distributed in the spectrum of aerosol sizes and that they are inactivated at the same rate. Importantly, we account for pre-symptomatic transmission of SARS-CoV-2. A newly infected individual has a 20% chance to be infectious before symptom onset and can start transmitting up to three days before symptom onset (58).

#### Grid search approach to define transmission pathways

The short- and long-range airborne transmission routes do not necessarily contribute at the same level in the transmission dynamics of every respiratory pathogen. Here, we explore five transmission pathways with varying contribution levels of the two transmission routes. These pathways are defined by choosing value pairs of (*β*_*e*,_ *β*_*c*_) that lead to similar transmission intensity in the ICU (see Appendix S3 for more details on the grid search approach).

#### Comparison of transmission pathways

For each transmission pathway, we simulate 500 epidemics. Resulting transmission dynamics are compared based on epidemic summary statistics: the time to the epidemic peak, the epidemic duration, the extinction probability (i.e. the probability that the transmission chain immediately dies out), the global SAR, and the SARs stratified either by infection source (close- or long-range transmission route) or by individual category (patient, medical staff, or paramedical staff). To compare the SARs resulting from the close- and long-range routes, we calculate the pseudo-median of the Wilcoxon rank sum test, which is the median of the differences between the two routes.

#### Intervention evaluation

We define and test seven strategies: (*i*) improved hand hygiene of HCWs, which limits the short-range transmission risk from infectious HCWs to patients, (*ii*) HCWs wear masks when they are in patient rooms or in interaction with patients and patients wear masks when they are symptomatic, which reduces infectious aerosol exhalation in patient rooms and the corridor and the risk of short-range transmission during patient-patient and patient-HCW interactions, (*iii*) universal mask wearing after the symptom onset of the index case, (*iv*) improved ventilation in rooms that can be visited by patients (patient rooms and corridor), (*v*) improved ventilation in rooms that are not accessible to patients (nursing station, medical office, medical and paramedical staff rooms), (*vi*) a mixed intervention that combines interventions (*ii*) and (i*v*), and (*vii*) a mixed intervention that combined (*ii*) and (*v*) (Table 2). We investigate, for each scenario, how individual and collective intervention measures affect transmission in ICUs and across individual categories. We assess the effectiveness of each intervention by dividing the pseudo-median of the difference in cumulative incidence from the Wilcoxon rank sum test by the median cumulative incidence when no intervention is in place. This allows us to evaluate the relative reduction in the 90-day incidence compared to the baseline scenario.

#### Sensitivity analysis

Most parameter values are fixed using estimates from the literature. While we present transmission dynamics using intermediate values for each parameter in the main analysis, we explore the influence of lower- and upper-bounds in the sensitivity analysis (Table 3). We notably test extreme values for the exhalation and inactivation rates in the absence of interventions. We also evaluate extreme values for the effectiveness of hand hygiene, masks, and ventilation.

The agent-based model is implemented using the Rcpp R package (59). The post-processing of simulated epidemics is implemented in R.

## Acknowledgments

The authors thank Prof Didier Guillemot, Dr Bich-Tram Huynh, and Dr Antoine Fraboulet for providing access to the Nods-Cov-2 data. The authors also thank Prof Djilalli Annane for his helpful comments on the organization of ICUs in France, Maximilien Lyonnais for his help on the updated version of the contact data processing pipeline, and Prof Didier Guillemot for the insightful discussions.

## Data availability

Interaction data from the Nods-Cov-2 project are not publicly available but can be made available from the corresponding author on reasonable request. All codes and synthetic data are available at https://github.com/MESuRS-Lab/aerosol-epi-model.

## Author Contributions

ML, LT, and LO designed the research. OG and ML performed the research with the support of QJL, JD, and GS. GS contributed to the analysis of the raw temporal networks. QJL generated the synthetic temporal networks. OG and JD generated the location reconstruction algorithm. OG, SK, LT, LO, and ML interpreted the results. OG and ML wrote the paper with the help of LT and LO. All authors reviewed the manuscript and took responsibility for the decision to submit for publication.

