## Supplementary material for "Accounting for the long-distance transmission route: an epidemiological model of airborne disease transmission in hospitals": SI Appendix

### Supplementary Materials

### 1. Close-proximity interactions data from the Nods-Cov-2 study

#### a. Description of the participants

We apply our simulation procedure to close-proximity interactions data of two adult French ICUs from the Nods-Cov-2 study (1), hereafter denoted as ICU1 and ICU2. Patients, HCWs, administration staff, logistic staff, and visitors who were present at the time of the Nods-Cov-2 study were equipped with wearable sensors recording interactions of less than 1.5 meters over a 36-hour period between April and June 2020. Recording was performed at the start of the COVID-19 pandemic in France, when the historical SARS-CoV-2 variant was circulating. Importantly, this period coincides with strong infection prevention and control measures in hospitals, notably visits from patient relatives were strongly restricted. Ward organization may not be completely representative of non-pandemic times.

In Supplementary Table 1, we detail the number of participants per ICU. Importantly, the number of paramedical staff is much higher in ICU1 compared to ICU2, despite both ICUs having a similar number of patients. We restrict our analysis to patients and HCWs which simplifies the reconstruction of individual locations. Finally, *n*=2 and *n*=2 participants had no interaction recorded over the study period in ICU1 and ICU2, respectively (Supplementary Table 2).

**Supplementary Table 1. Number of participants in the two adult intensive care units of the NODS-CoV-2 study.** In the main analysis, we focused on patients and healthcare workers, thus we excluded administration staff, logistics staff, visitors, and participants of unknown category in the main analysis.

| | | **ICU1** | **ICU2** |
|---|---|---|---|
| **Patients** | | 18 | 17 |
| **Healthcare workers** | | | |
| | Medical staff | 30 | 29 |
| | Paramedical staff | 118 | 46 |
| **Other** | | | |
| | Administration | 10 | 6 |
| | Logistics | 1 | 10 |
| | Visitor | 1 | 0 |
| | Unknown | 16 | 4 |
| **Total** | | 194 | 112 |

**Supplementary Table 2. Number of individuals without interaction over the study period.**

| | **ICU1** | **ICU2** |
|---|---|---|
| Medical staff | 2 | 1 |
| Paramedical staff | 0 | 1 |

##### b. Comparison of the observed and synthetic temporal networks

Due to the short time window of the close-proximity interactions recorded as of the Nods-Cov-2 study, the number of patients is too small to simulate realistic outbreaks over 90 days. Thus, we augment the number of patients to reproduce realistic hospitalization stays in French ICUs. We then use a previously published algorithm (2) to generate synthetic temporal networks between HCWs and patients (observed and augmented). This algorithm stochastically generates contacts between individuals using the empirical distribution of contacts between pairs of individual categories (HCW-HCW, patient-patient, and HCW-patient) per hour of the day. This algorithm allows the reconstruction of contacts for individuals that do not have contacts in the observed temporal networks and the augmented patients. Finally, we truncate interactions to match the schedule of the HCWs and hospitalization stays of patients.

In Supplementary Figure 1, we compare (*i*) the dynamics of the number of unique contacts per pairs of individual categories and (*ii*) the distribution of contact durations in the observed temporal network and 10 examples of synthetic temporal networks. Synthetic temporal networks reproduce well interactions between individuals as well as their duration.

In Supplementary Figure 2, we compare network properties of the observed temporal network and the synthetic temporal network used in the main analysis. This comparison highlights that the algorithm does not necessarily reconstruct faithfully the same properties across observed networks. Here, we obtain consistent degrees, densities and efficiencies for ICU1, while transitivity (density of triangles) is best reproduced in ICU2.

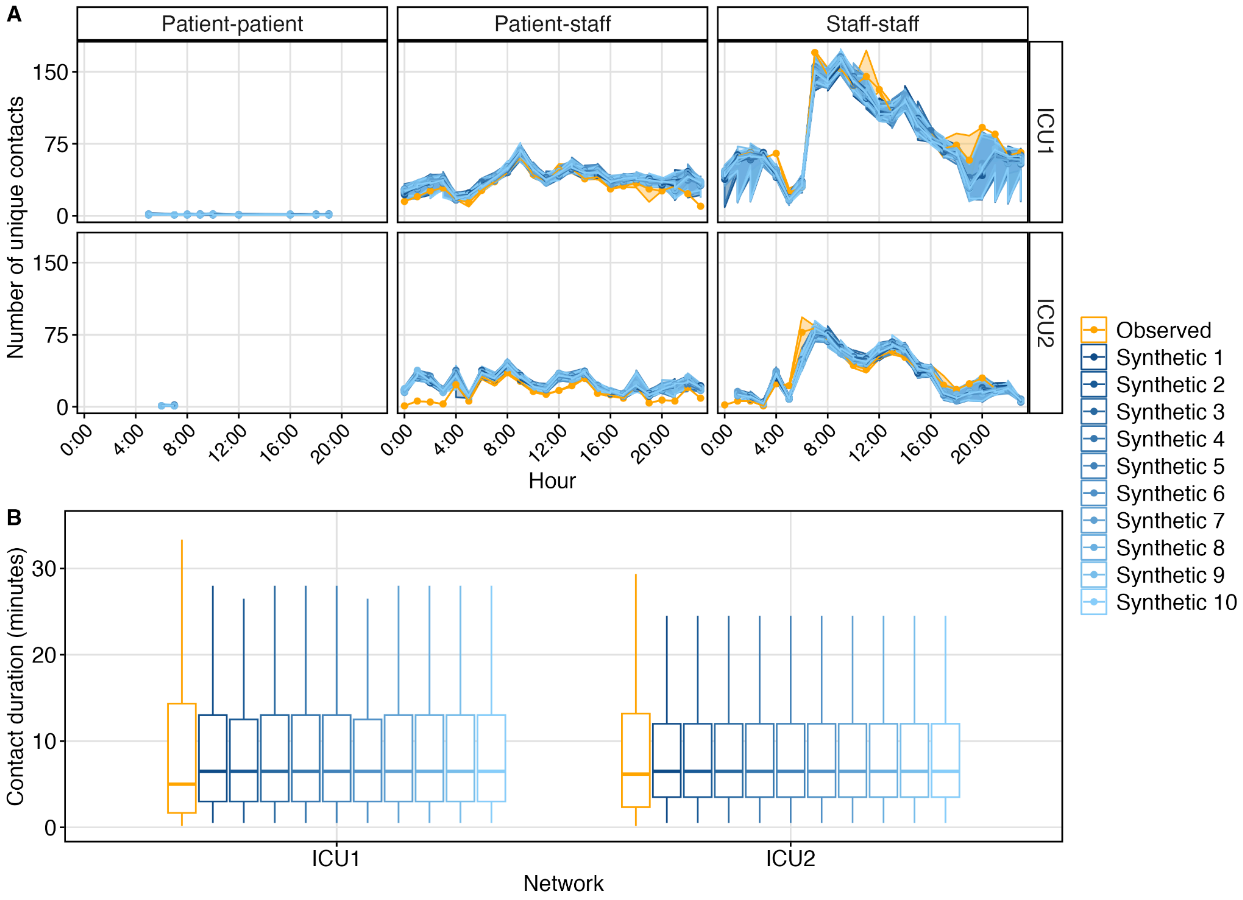

**Supplementary Figure 1. Contact patterns of observed and synthetic temporal networks in ICU1 and ICU2.** The temporal network reconstruction algorithm is stochastic. We provide here a comparison between ten examples of reconstructed temporal networks (shades of blue) and the corresponding observed temporal network (orange). (**A**) Number of unique contacts per hour in the observed and synthetic temporal networks stratified by pairs of individuals. The solid line corresponds to the median number of unique contacts and the area to the 25% and 75% percentiles. (**B**) Distribution of contact durations (in minutes) in the observed and synthetic temporal networks.

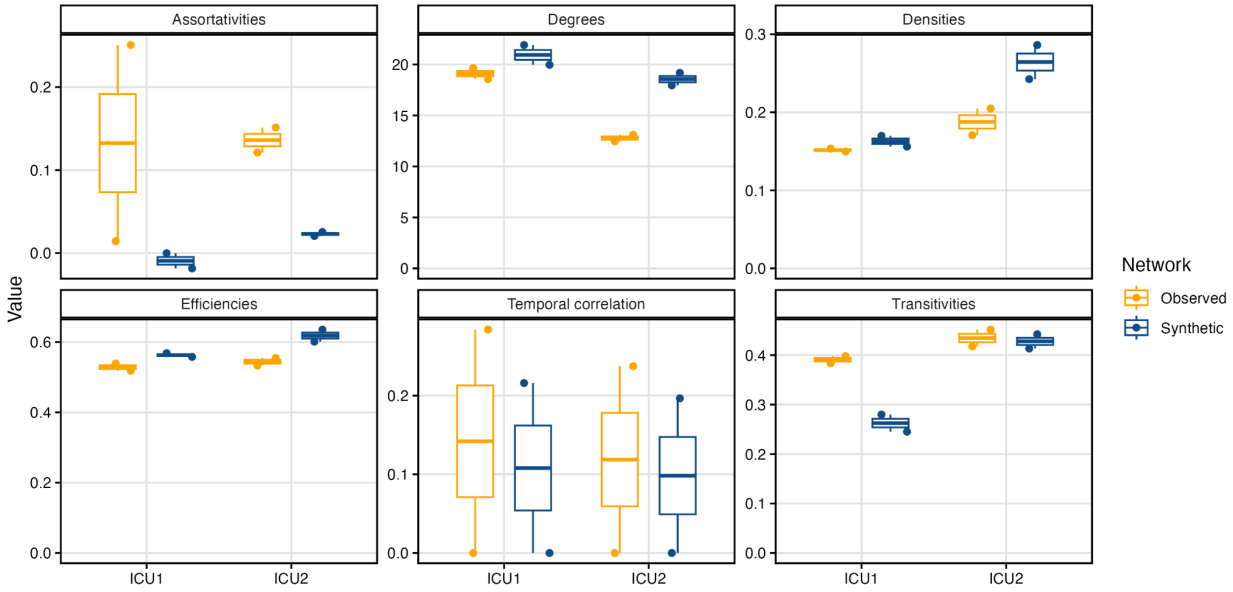

**Supplementary Figure 2. Comparison of network metrics for the observed and synthetic temporal networks in ICU1 and ICU2.** We compare network properties of the observed temporal networks (in orange) and the synthetic temporal networks (in blue) used in the epidemic simulation study. The two points correspond to the two days of recording in the observed networks and the first two days of the reconstruction in the synthetic networks. The assortativity corresponds to the degree to which similar vertices (i.e. vertices of the same category among patients, medical staff, paramedical staff) tend to connect to each other. The degree is the mean number of contacts. The density is the proportion of edges among all potential edges in the network. The efficiency is the average of the inverse distances between all pairs of vertices. The temporal correlation is the average probability that between two subsequent days, an individual maintains the same number of unique contacts with the same individuals. The transitivity is the probability that adjacent vertices of a vertex are connected.

#### 2. Location reconstruction

##### a. Algorithm to reconstruct individual locations based on their category and interactions

Our transmission model describes infection due to the exposure to infectious aerosols in rooms that are visited by patients and HCWs. Due to the absence of location data at fine temporal scales, we develop a location reconstruction procedure that reconstructs location of individuals in a minimal ward configuration. This minimal ward is composed of patient rooms, a nursing station, a medical office, a room for the medical staff, a room for the paramedical staff, and a corridor that connects all the rooms. This configuration is applied to both ICUs and does not aim to accurately describe the specific contexts of ICU1 and ICU2. Here, we intend to develop a generic tool that relies on few assumptions, and remains flexible enough to apply to different contexts. Nevertheless, we adapt the volume of the different rooms so that the minimal volume per individual is the same in both ICUs.

The location reconstruction procedure is divided into three steps, as follows:

1. Location assignment during close-proximity interactions (Supplementary Figure 3B): locations depend on the individual's category and the composition of the interaction cluster they belong to. Importantly, individuals within the same cluster are not necessarily in interaction with all the other members of the cluster. For example, multiple HCWs may be in contact with the same patient but not on the same side of the patient's bed, thus separated by more than 1.5 meters.
2. Individual location smoothing (Supplementary Figure 3C): in this step, we assume that an individual remains in the same room if there is a gap of less than five minutes in a sequence within the same room.
3. Location assignment when individuals are not in interaction (Supplementary Figure 3D): HCWs with no interaction for more than 30 minutes are assumed to be out of the ICU, while patients remain in their room.

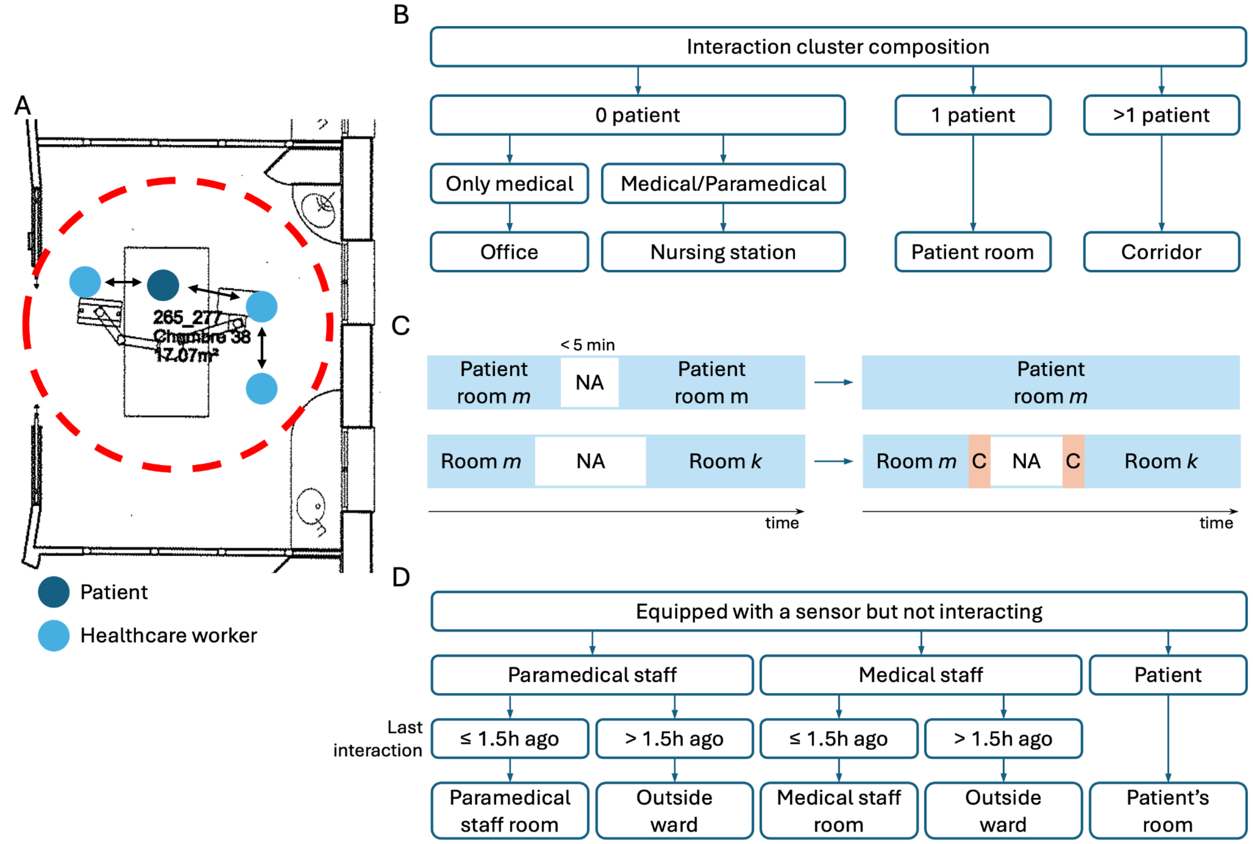

**Supplementary Figure 3. Location reconstruction procedure.** (**A**) Example of an interaction cluster. (**B**) First step of the location reconstruction procedure. In this step, we reconstructed the location of individuals when they are interacting. (**C**) Second step of the location reconstruction procedure which consists in smoothing spatial trajectories. (**D**) In the third step of the location reconstruction procedure, we reconstructed the location of individuals when they are within the ward but not interacting. In a sensitivity analysis, we explored how different cut-offs (30 min, 1h, and 1.5h) of the time spent without interactions before considering individuals to leave the ward at not being exposed to aerosols suspended in the air of the different ward rooms.

##### b. Comparison of reconstructed locations between synthetic and observed temporal networks

As presented in Supplementary Figure 2, some network properties are less conserved between the observed and synthetic temporal networks, notably network transitivity for ICU1 and network density for ICU2. This has a direct implication on the reconstruction of individual locations illustrated in Supplementary Figure 4. In synthetic networks, there is a slight difference in the cumulative time spent by medical staff, paramedical staff, and patients in the different rooms of the minimal ward compared to what is observed in the Nods-CoV-2 study. Notably, patients spend more time in the corridor (i.e. in interaction with other patients), compared to the observed data. Nevertheless, simulated individual movements are consistent and our model is able to generate aerosol dynamics in the different rooms through the movement of infectious individuals.

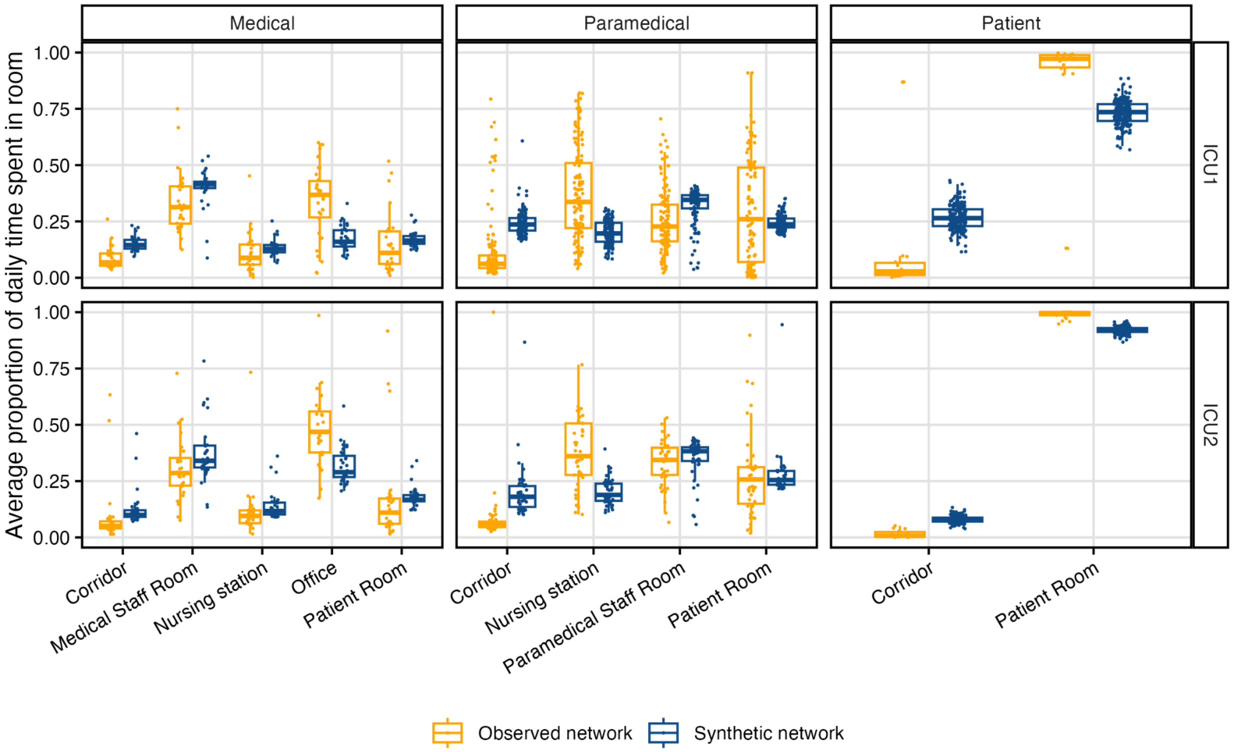

**Supplementary Figure 4. Comparison of the proportion of daily time spent in the rooms of the ward between the observed and synthetic temporal networks.** For each individual (one circle), we compute the average proportion of the time spent in the different rooms of the ward over their days of presence in the ward. For example, the proportion is averaged over the stay lengths of patients, while it is averaged over the working days for HCWs. This procedure is applied to the observed (in orange) and synthetic (in blue) networks.

#### 3. Simulation study

##### a. Grid search procedure to select parameter values for the transmission scenarios

We explore value pairs of $(\beta_e, \beta_c)$ using a grid search approach and select five value pairs leading to similar secondary attack rates (SAR), defined as the proportion of susceptible individuals that got infected during the 90-day simulation period after the introduction of the index case (Supplementary Figure 5). Selected values are listed in Table 1.

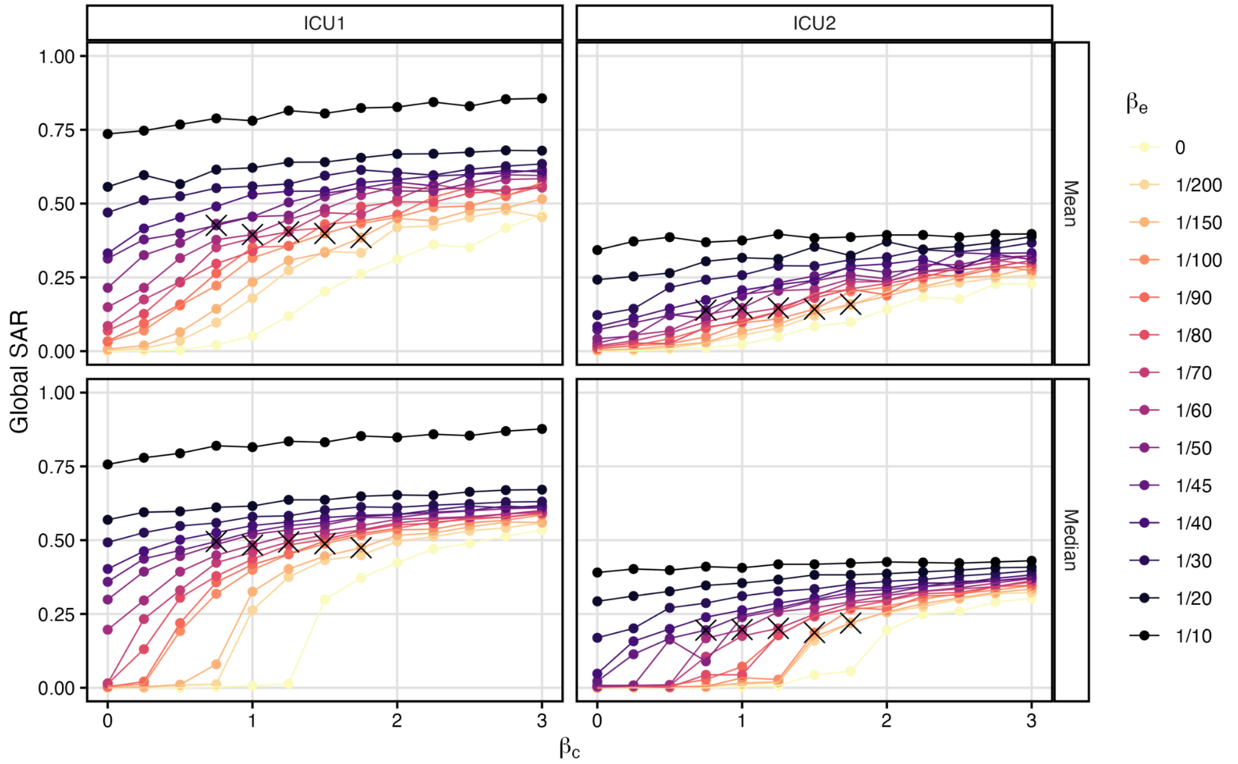

**Supplementary Figure 5. Grid search procedure.** For each network and value pair of ($\beta_e$, $\beta_c$), we simulate *n*=100 epidemics and report the median and average secondary attack rates (SAR). This grid search approach allows us to select five value pairs of ($\beta_e$, $\beta_c$) depicted with black crosses that represent the five scenarios of transmission explored in the main analysis. These value pairs are chosen so that SARs are similar within each ICU network.

##### b. Decomposition of short-range transmission in the baseline scenario without intervention

In all transmission pathways, most of the short-range transmission events occur between HCWs, followed by transmission from HCWs to patients, from patient to HCWs, and finally transmission between patients (Supplementary Figure 6). This is the case in both ICUs which highlights the central role of HCWs in the dynamics of healthcare-acquired infections.

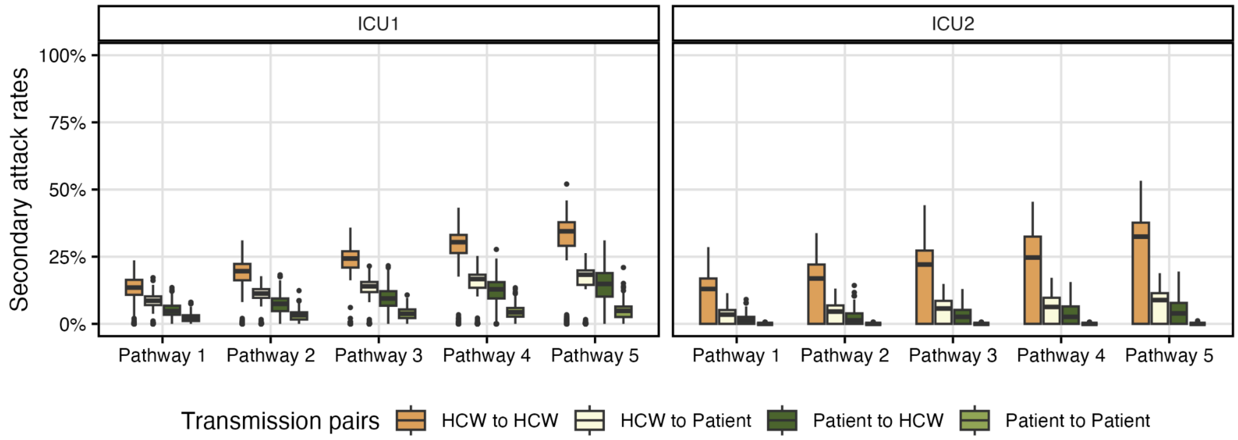

**Supplementary Figure 6. Transmission pairs during short-distance transmission events.** We report the secondary attack rates among patients or HCWs depending on the category of their infector during short-distance infections. In all transmission scenarios, the highest secondary attack rates are due to infections between HCWs, followed by infections from HCWs to patients, then patients to HCWs and finally patients to patients that is anecdotical in ICU2. As expected, when the short-distance route is predominant, SARs increase for all pairs.

Pathway 1: $\beta_e$ = 1/45 new cases per infectious aerosol per day and $\beta_c$ = 0.75 new case per contact per day; Pathway 2: $\beta_e$ = 1/60 and $\beta_c$= 1.00; Pathway 3: $\beta_e$ = 1/70 and $\beta_c$ = 1.25; Pathway 4: $\beta_e$ = 1/100 and $\beta_c$ = 1.50; and Pathway 5: $\beta_e$ = 1/150 and $\beta_c$ = 1.75.

##### c. Comparison of intervention effectiveness

In Supplementary Figure 7, we present intervention effectiveness in patients, in paramedical staff, and in medical staff. We also report effectiveness among HCWs by pooling paramedical and medical staff, as well as the overall effectiveness, when considering all individuals. Strikingly, only two interventions, universal masking and ventilation in rooms that are inaccessible to patients, reduce transmission among HCWs as much as among patients. Given that HCWs are a major source of infection for patients (Supplementary Figure 6), interventions should also target HCWs. Overall, universal masking is the most effective intervention in both ICUs and across all transmission pathways (Supplementary Figure 8). Improving ventilation in rooms that are inaccessible to patients indirectly contributes to the reduction of short-range transmission by HCWs (Supplementary Figure 9). This is the case in Pathway 1 and Pathway 2 where the long-range transmission is predominant.

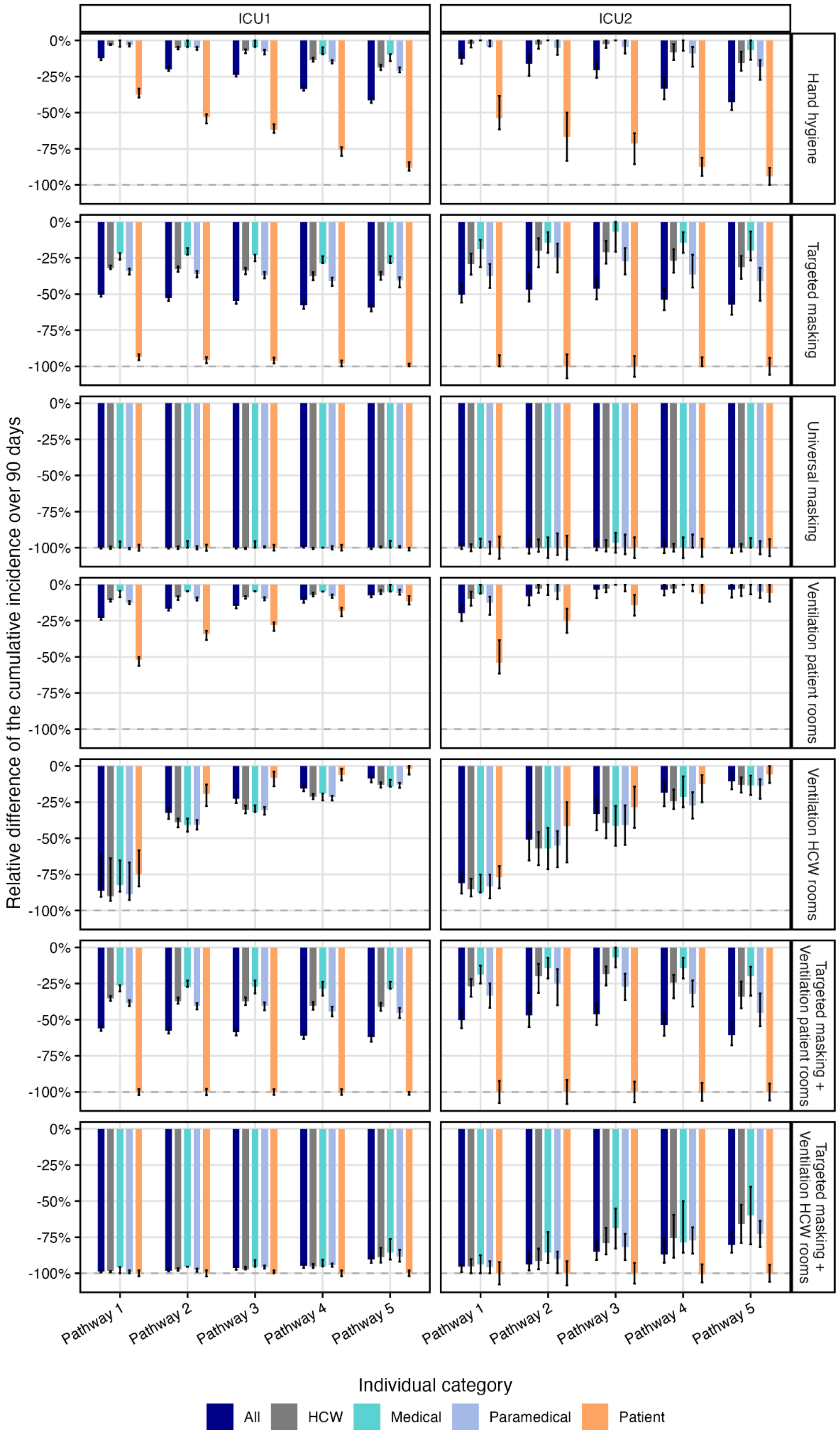

**Supplementary Figure 7. Relative difference of the cumulative incidence over 90 days compared to the baseline scenario without intervention and stratified by individual category.** Relative difference of the 90-day cumulative incidence among a specific category of individuals between the intervention scenario and the baseline scenario (with no intervention). Negative values indicate a reduction in the cumulative incidence when an intervention is in place. Relative difference estimates and error bars are derived from the pseudo-median and the 95% CI of Wilcoxon tests, respectively. Reductions stronger than 100% are allowed since we used the median cumulative incidence in the baseline scenario as the reference and the 95% CI bounds of the difference can exceed this median.

Pathway 1: $\beta_e$ = 1/45 new cases per infectious aerosol per day and $\beta_c$ = 0.75 new case per contact per day; Pathway 2: $\beta_e$ = 1/60 and $\beta_c$= 1.00; Pathway 3: $\beta_e$ = 1/70 and $\beta_c$ = 1.25; Pathway 4: $\beta_e$ = 1/100 and $\beta_c$ = 1.50; and Pathway 5: $\beta_e$ = 1/150 and $\beta_c$ = 1.75.

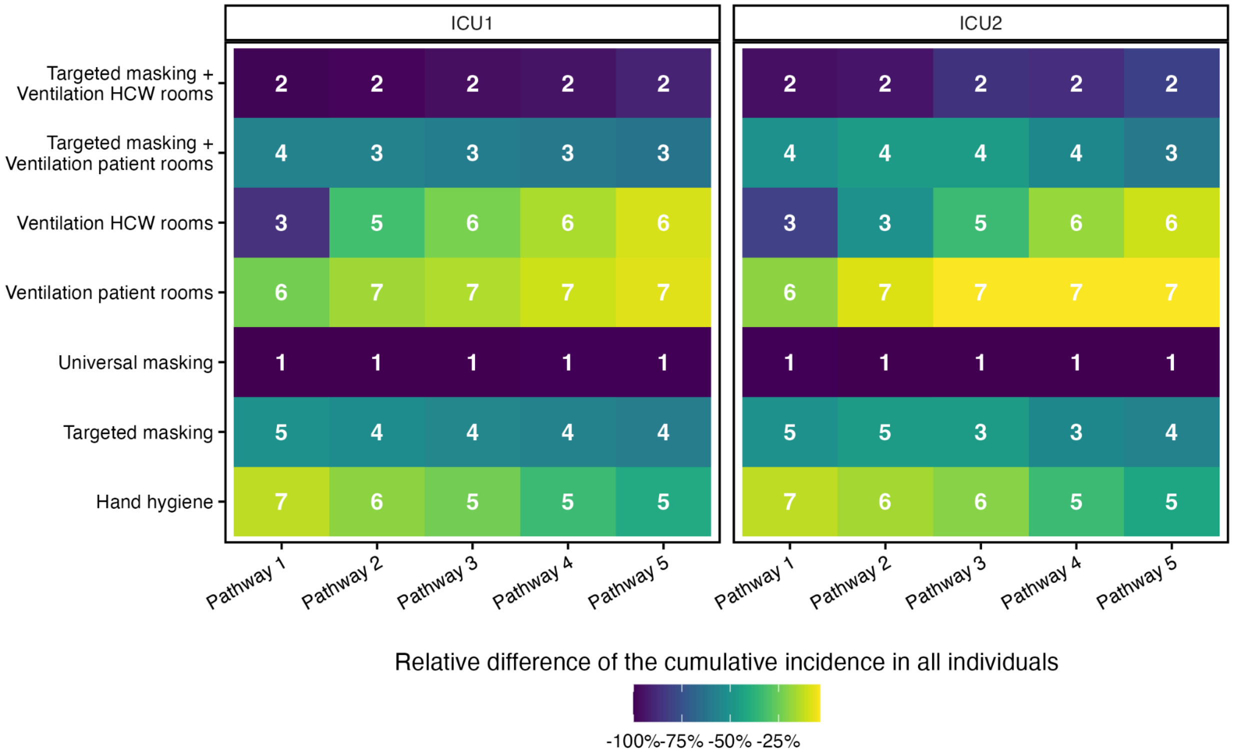

**Supplementary Figure 8. Ranking of the most effective intervention in reducing the overall cumulative incidence over 90 days.** For each combination of ICU and transmission pathway, we rank the effectiveness of interventions based on the relative difference in the cumulative incidence over 90 days compared to the scenario without intervention. Universal masking emerges as the most effective intervention in both ICUs and across all transmission pathways.

Pathway 1: $\beta_e$ = 1/45 new cases per infectious aerosol per day and $\beta_c$ = 0.75 new case per contact per day; Pathway 2: $\beta_e$ = 1/60 and $\beta_c$= 1.00; Pathway 3: $\beta_e$ = 1/70 and $\beta_c$ = 1.25; Pathway 4: $\beta_e$ = 1/100 and $\beta_c$ = 1.50; and Pathway 5: $\beta_e$ = 1/150 and $\beta_c$ = 1.75.

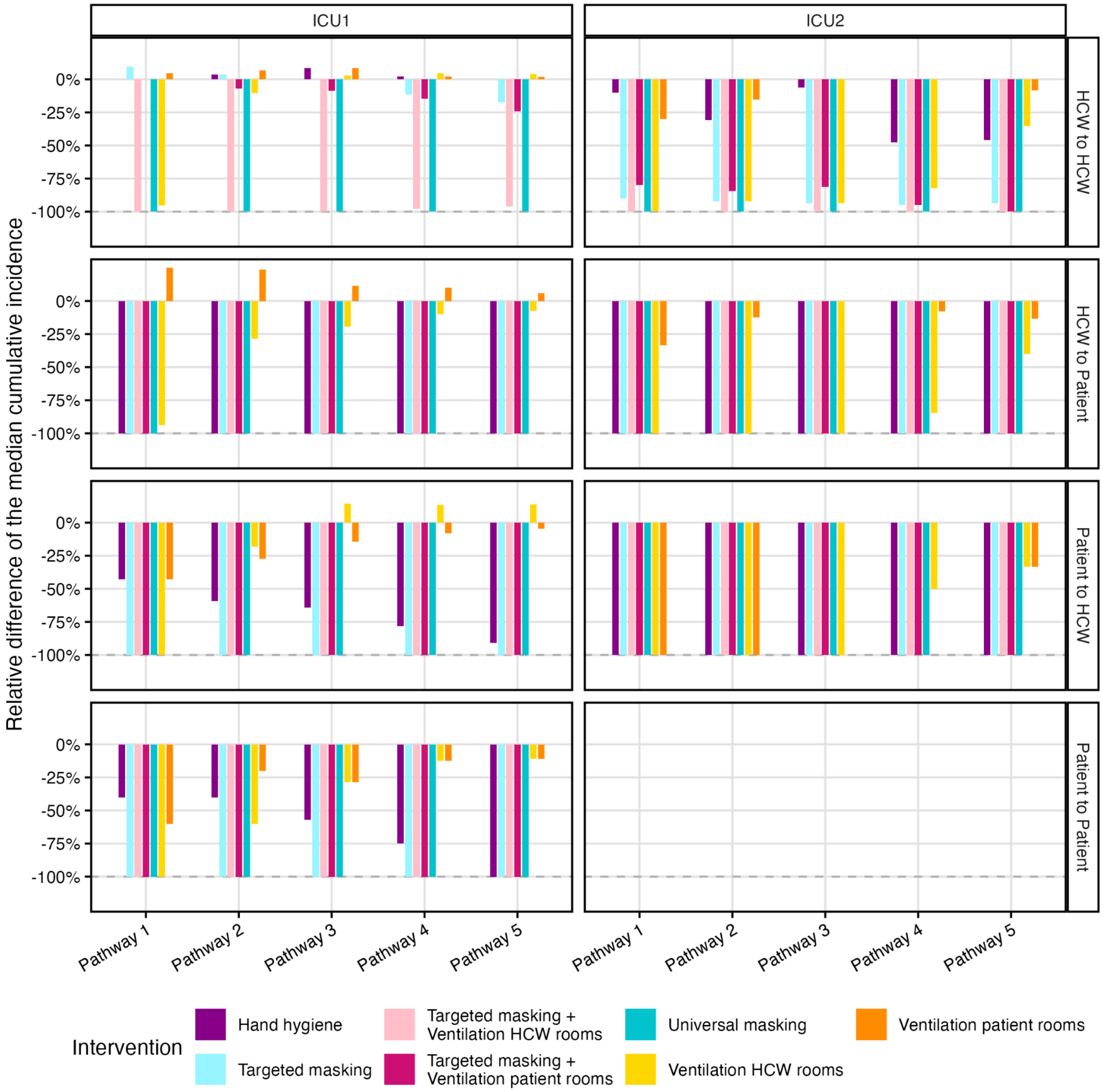

**Supplementary Figure 9. Relative difference of the median cumulative incidence for each transmission pair compared to the baseline scenario.** Barplots correspond to the relative difference of the median cumulative incidence in the intervention scenario compared to the baseline scenario. In ICU2, the median of the patient-to-patient cumulative incidence is equal to 0 for all transmission scenarios with and without interventions. The relative difference cannot be computed in this specific case.

Pathway 1: $\beta_e$ = 1/45 new cases per infectious aerosol per day and $\beta_c$ = 0.75 new case per contact per day; Pathway 2: $\beta_e$ = 1/60 and $\beta_c$= 1.00; Pathway 3: $\beta_e$ = 1/70 and $\beta_c$ = 1.25; Pathway 4: $\beta_e$ = 1/100 and $\beta_c$ = 1.50; and Pathway 5: $\beta_e$ = 1/150 and $\beta_c$ = 1.75.

##### d. Sensitivity analyses in scenarios with interventions

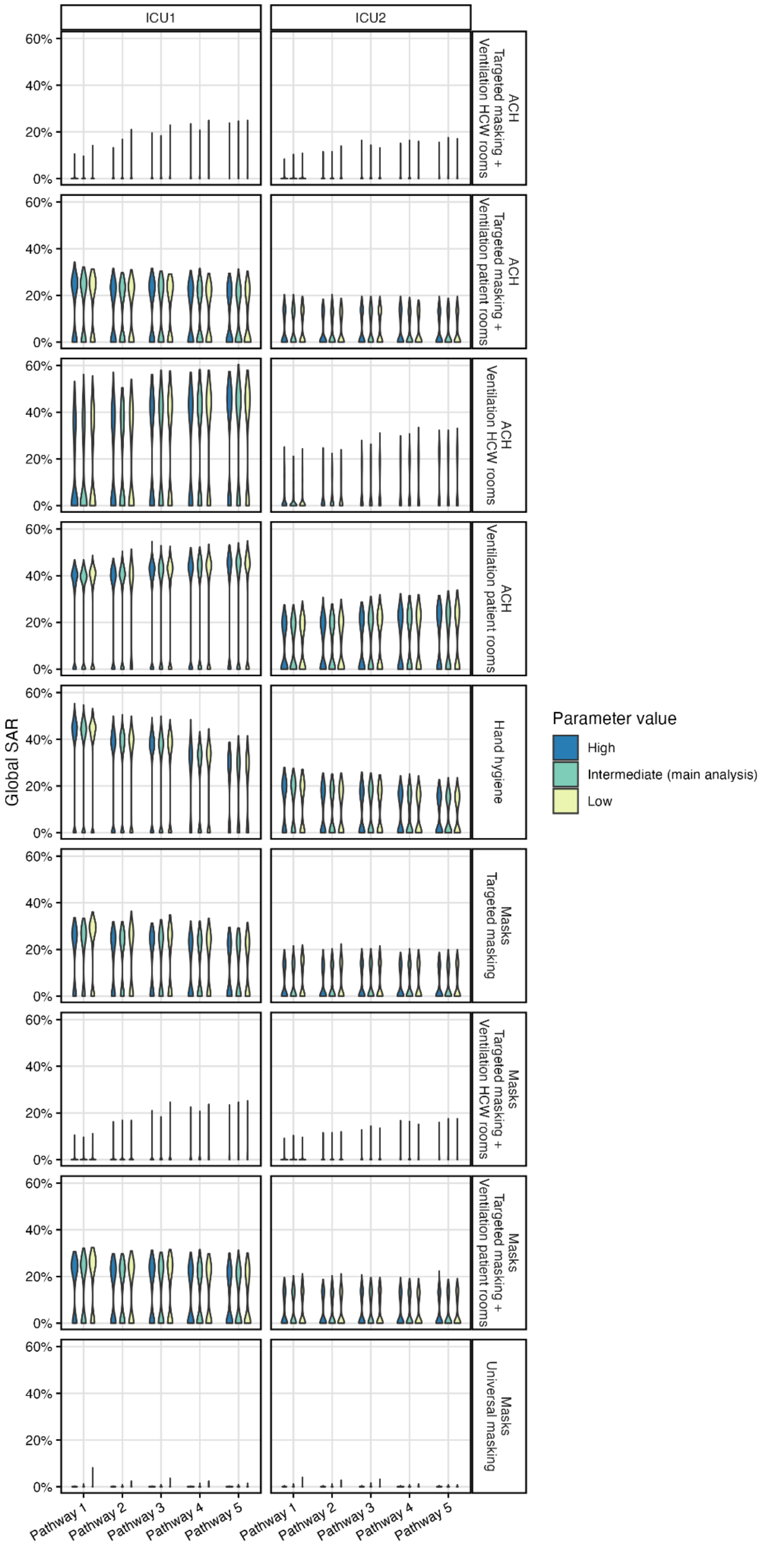

**Supplementary Figure 10. Distributions of secondary attack rates in the sensitivity analyses related to intervention effectiveness.** We tested low, intermediate, and high values of key parameters governing the effectiveness of the different interventions (in rows) in a univariate sensitivity analysis across the two ICU networks and the five transmission scenarios. Parameter values are detailed in Table 3.

Pathway 1: $\beta_e$ = 1/45 new cases per infectious aerosol per day and $\beta_c$ = 0.75 new case per contact per day; Pathway 2: $\beta_e$ = 1/60 and $\beta_c$= 1.00; Pathway 3: $\beta_e$ = 1/70 and $\beta_c$ = 1.25; Pathway 4: $\beta_e$ = 1/100 and $\beta_c$ = 1.50; and Pathway 5: $\beta_e$ = 1/150 and $\beta_c$ = 1.75.

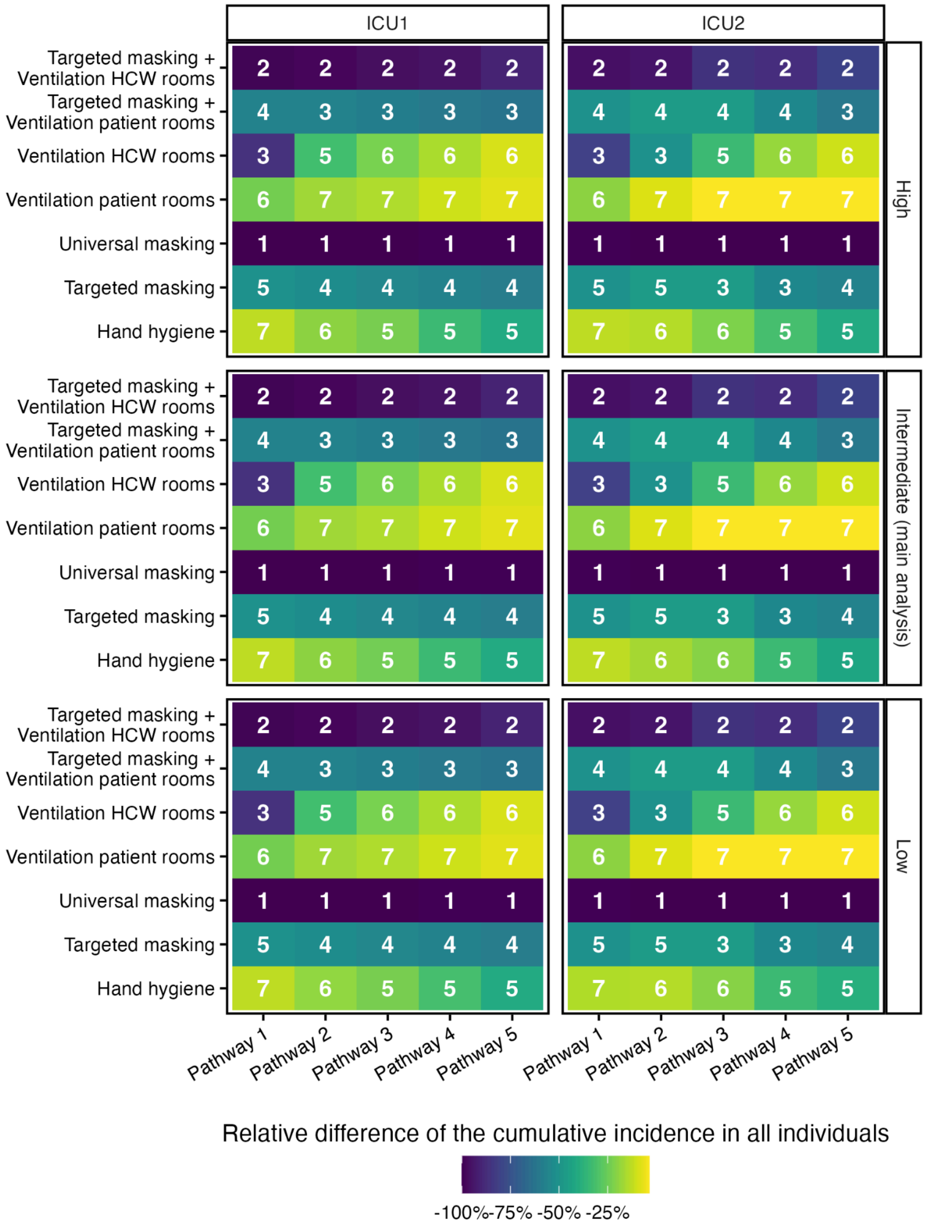

**Supplementary Figure 11. Ranking of the most effective interventions in reducing the overall cumulative incidence over 90 days for varying levels of efficacy of hand hygiene.**

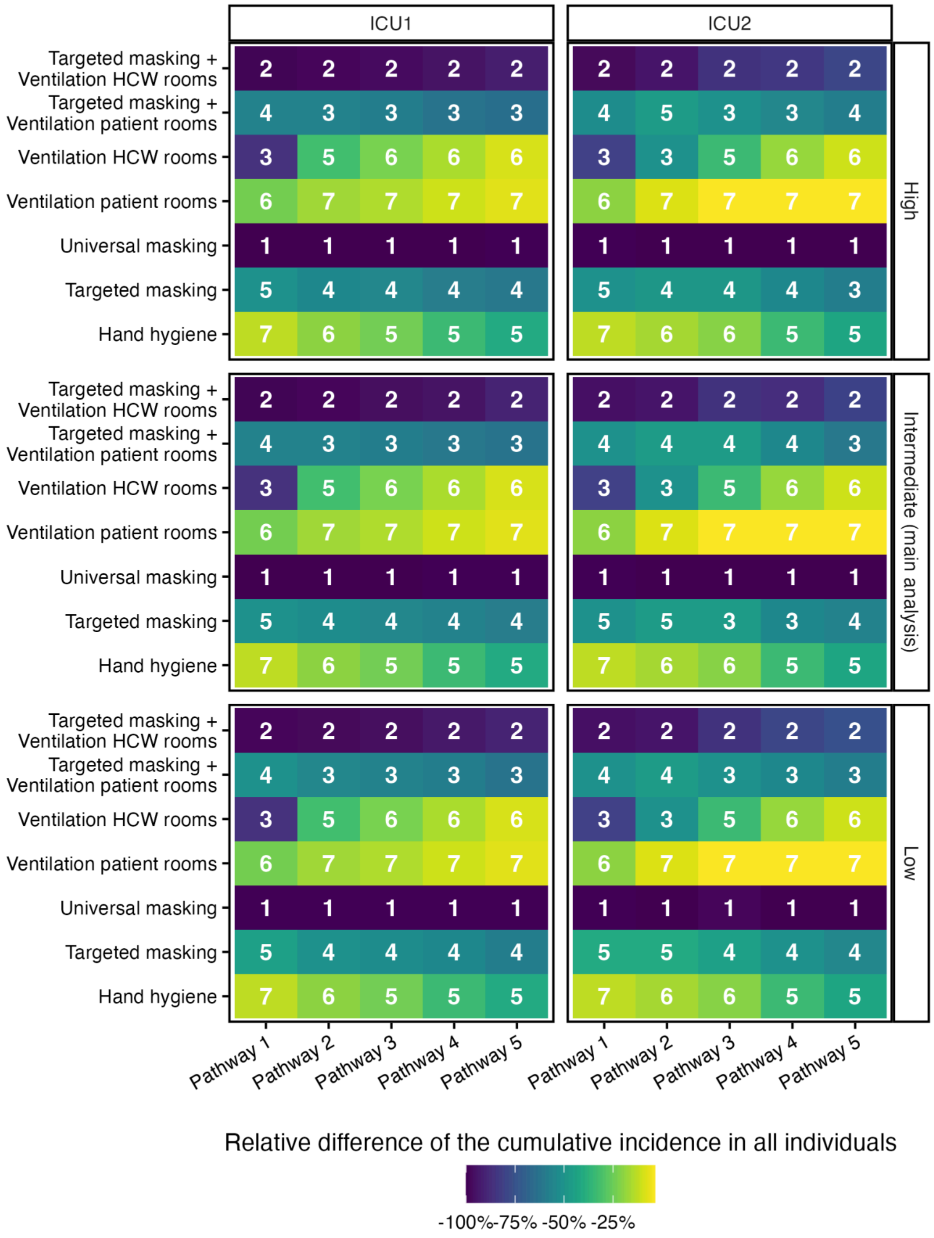

**Supplementary Figure 12. Ranking of the most effective interventions in reducing the overall cumulative incidence over 90 days for varying levels of efficacy of mask wearing.**

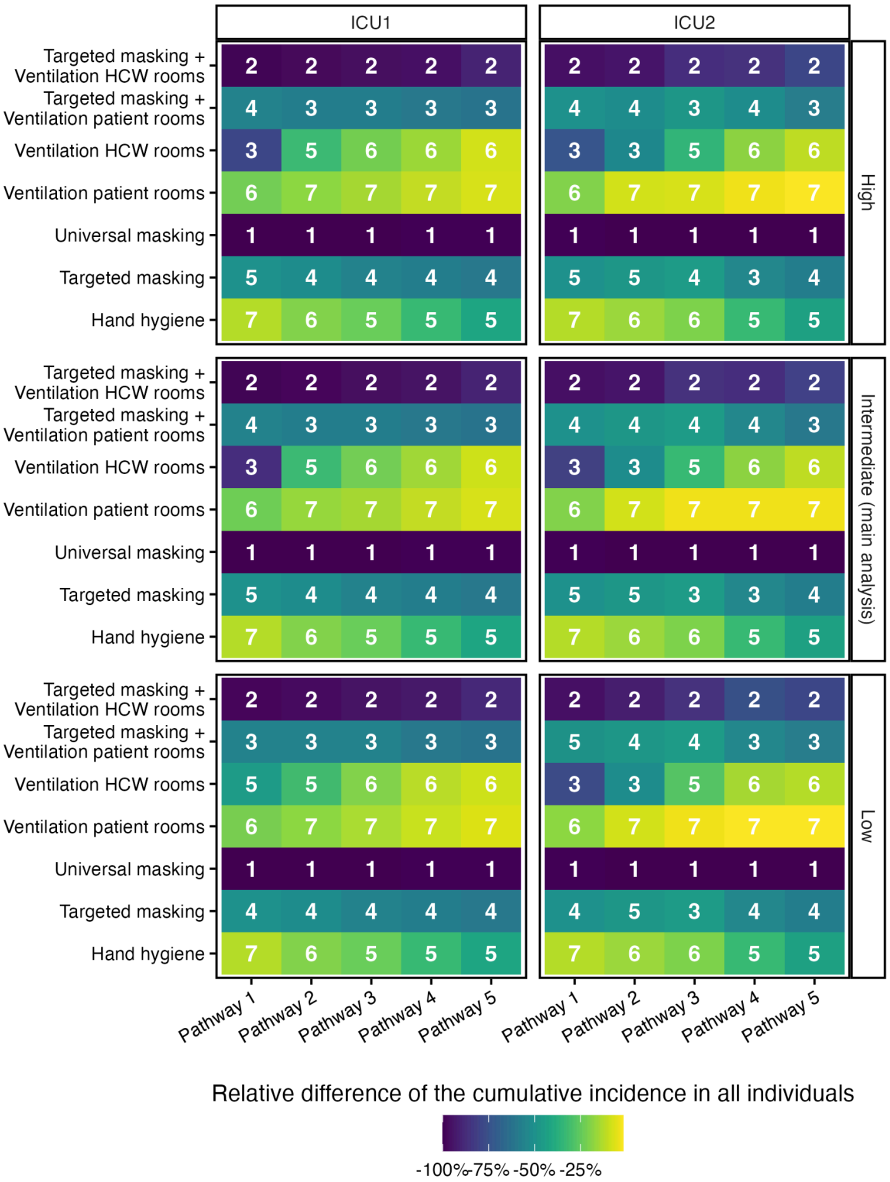

**Supplementary Figure 12. Ranking of the most effective interventions in reducing the overall cumulative incidence over 90 days for varying levels of efficacy of ventilation.**
